# Clinical and Non-clinical Proof of Concept Supporting the Development of RJX As an Adjunct to Standard of Care Against Severe COVID-19

**DOI:** 10.1101/2022.02.12.22270748

**Authors:** Fatih M. Uckun, Muhammad Saeed, Mustafa Awili, Ibrahim H. Ozercan, Sanjive Qazi, Cynthia Lee, Adeel Shibli, Alan W. Skolnick, Alonso Prusmack, Joseph Varon, Cesar I.P. Barrera, Cemal Orhan, Michael Volk, Kazim Sahin

## Abstract

**Background:** The identification of effective strategies capable of reducing the case mortality rate of high-risk COVID-19 is an urgent and unmet medical need. We recently reported the clinical safety profile of RJX, a well-defined intravenous GMP-grade pharmaceutical formulation of anti-oxidant and anti-inflammatory vitamins as active ingredients, in a Phase 1 study in healthy volunteers (ClinicalTrials.gov Identifier: NCT03680105) (Uckun et al., Front. Pharmacol. 11, 594321. 10.3389/fphar.2020.594321). Here we report data from a pilot clinical study (RPI-015) which examined the safety, tolerability, and feasibility of using RJX in combination with clinical standard of care (SOC) in hospitalized COVID-19 patients with pneumonia (ClinicalTrials.gov Identifier: NCT04708340). In addition to our early clinical proof of concept (POC) data, we also present non-clinical POC from a mouse model of CRS and ARDS that informed the design of the reported clinical study.

**Methods:** 13 patients, who were hospitalized with COVID-19 pneumonia and abnormally elevated serum inflammatory biomarkers markers ≥3 months prior to the identification of the first confirmed U.S case of the Omicron variant, were treated with IV RJX (daily x 7 days) plus SOC. Non-clinical POC study examined the ability of RJX plus dexamethasone (DEX) to improve the survival outcome in the lipopolysaccharide (LPS)-Galactosamine (GalN) mouse model of fatal cytokine release syndrome (CRS), sepsis and acute respiratory distress syndrome (ARDS).

**Findings:** In the Phase 1 clinical study, none of the 13 patients developed a treatment-related DLT, SAE, or Grade 3-5 AEs. Nine (9) of the 12 evaluable patients, including 3 patients with hypoxemic respiratory failure, showed rapid clinical recovery. In the non-clinical POC study in LPS-GalN challenged mice, the combination of RJX plus DEX was more effective than RJX alone or DEX alone, reversed the CRS as well as inflammatory tissue damage in the lungs and liver, and improved the survival outcome. Taken together, these findings provide the early clinical and non-clinical POC for the development of RJX as an adjunct to the SOC in the multi-modality management of high-risk COVID-19.

## Introduction

COVID-19 has a high fatality rate in elderly and high-risk populations with comorbidities, such as hypertension, diabetes, obesity, immunodeficiencies and hematologic cancers [1-12]. Several pro-inflammatory cytokines, including interleukin-1 (IL-1), interleukin-6 (IL6), tumor necrosis factor-alpha (TNF-α), and transforming growth factor-beta (TGF-β), contribute to the development of a potentially fatal severe systemic inflammation in high-risk COVD-19 patients, which is known as cytokine release syndrome (CRS) and is often associated with acute lung injury (ALI) leading to ARDS and potentially multi-organ dysfunction (MOD) in rapid progression [13-23]. A ∼40% likelihood of progression to severe-critical disease and ARDS within 10 days after symptom onset has been reported for high-risk COVID-19 patients [24-26] The identification of effective strategies capable of preventing the development of potentially fatal viral sepsis, cytokine release syndrome (CRS) and ARDS in high-risk COVID-19 patients, is an urgent and unmet medical need.

Rejuveinix (RJX) is a formulation of several anti-inflammatory and anti-oxidant vitamins [26--34]. RJX exhibited promising single-agent anti-inflammatory activity in a mouse model of fatal CRS and ARDS [26]. Preliminary evidence suggested that it may effectively reverse CRS and ARDS if administered after the onset of ALI in LPS-GalN challenged mice [35]. Further, RJX showed a very favorable clinical safety and pharmacokinetics (PK) profile in a recently completed randomized, double-blind, placebo-controlled Phase I ascending dose-escalation study in healthy volunteers (ClinicalTrials.gov Identifier: NCT03680105) [26].

Here we report data from a pilot clinical study which examined the safety, tolerability, and feasibility of using RJX in combination with clinical standard of care (SOC), including low dose Dexamethasone (DEX) (or Solumedrol) in hospitalized COVID-19 patients with pneumonia (ClinicalTrials.gov Identifier: NCT04708340). All patients were enrolled ≥3 months prior to the identification of the first confirmed U.S case of the Omicron variant. In addition to our early clinical proof of concept (POC) data, we also present non-clinical POC from a mouse model of CRS and ARDS that informed the design of the reported pilot clinical study. In the clinical study, no patient developed a treatment-related DLT, SAE, or Grade 3-5 AEs. Seventy-five % of patients showed rapid clinical recovery with resolution of the hyperinflammatory response and oxygen therapy requirements. In the non-clinical POC study in LPS-GalN challenged mice, the combination of RJX plus DEX was more effective than RJX alone or DEX alone, reversed the sepsis-associated CRS as well as inflammatory tissue damage in the lungs and liver, and improved the survival outcome. Taken together, these findings provide the early clinical and non-clinical POC for the development of RJX as an adjunct to the SOC in the multi-modality management of severe COVID-19 patients with viral sepsis and indicate its clinical impact potential.

## Materials and methods

### Rejuveinix (RJX)

RJX is a GMP-grade pharmaceutical composition of anti-inflammatory and anti-oxidant vitamins (Table S1 and Supplemental Methods) [26,36].

### Study Design and Eligibility Criteria

The safety, tolerability, and feasibility of using RJX in combination with clinical standard of care for hospitalized COVID-19 patients were evaluated in an open-label pilot study that was Part 1 of an ongoing double-blind, randomized, Phase 1/2 Study (ClinicalTrials.gov Identifier: NCT04708340). 13 patients who were hospitalized with COVID-19 pneumonia and abnormally elevated serum inflammatory biomarkers markers were with IV RJX (daily x 7 days) plus SOC. All patients were enrolled between April 5, 2021, and September 1, 2021, prior to the identification of the first case of the Omicron variant on December 1, 2021. The follow-up was complete through December 9, 2021. The primary goal of the study was to evaluate the safety and tolerability of RJX plus SOC in hospitalized severe COVID-19 patients. The secondary goal was to evaluate the clinical efficacy of RJX plus SOC in hospitalized severe COVID-19 patients. The Eligibility Criteria are detailed in the clinical protocol synopsis posted at ClinicalTrials.gov ((ClinicalTrials.gov Identifier: NCT04708340). Patients received daily 40 min (±10 min) infusions of 20 mL RJX mixed with 100 mL normal saline (total volume = 120 mL) intravenously plus SOC for 7 consecutive days. Some patients who no longer required inpatient care due to rapid improvement of their medical condition were discharged before day 7 and consequently received fewer than 7 treatments of RJX. SOC was provided until the patient was discharged. AEs were graded according to National Cancer Institute (NCI) Common Terminology Criteria for Adverse Events Version 5.0 (NCI CTCAE (v5.0). An 8-point ordinal scale was used as an assessment of the clinical status at the first assessment of a given study day (Supplemental Methods).

### Study Conduct

The study was performed under IND149585 at the following 4 centers in the US as an open-label study sponsored by Reven Pharmaceuticals: (1) Memorial Hermann Memorial City Medical Center, Houston, Texas; (2) Christus Health Santa Rosa Hospital, New Braunfels, Texas; (3) LFROS Research and United Memorial Medical Center, Houston, Texas; (4) PRX Research and Dallas Regional Medical Center, Dallas, Texas.

### Patient Disposition and Patient Characteristics

Twenty-one patients were screened for the clinical study. The details of patient disposition are shown in Figure S1. There were 8 screen failures. Thirteen hospitalized adult patients with severe or critical COVID-19, including 8 males and 5 females with a median age of 46 years (Range: 24-72 years) of whom 11 were White (Hispanic or Latino) and 2 were Asian (not Hispanic or Latino) were enrolled in one of the 2 cohorts of the clinical study within 0-11 days (Median: 4 days; Mean ± SE=4.5±1.0) days after initial COVID-19 diagnosis (Table S2). All 13 patients had imaging evidence of viral pneumonia and they were hypoxic requiring oxygen therapy. All patients were symptomatic with cough and shortness of breath. Nine patients had a fever as well. The mean and median BMI values were 35.6±1.9 and 35.1, respectively (Range: 23.8-49.4). Ten patients were obese with BMI values ranging from 32.1 to 49.4 and 2 patients were overweight with BMI values of 28.3 and 29.3, respectively. Five patients had HTN, 3 patients had diabetes, and 3 patients had pulmonary co-morbidities (asthma or COPD). Only one patient was vaccinated. All 13 patients had elevated serum CRP values ranging from 42.4 to 297.6 (Median: 64.3; Mean ± SE = 91.8±19.1). The ALC values ranged from 0.3 to 1.4 (Median: 0.9; Mean ± SE= 0.8±0.1). Eleven patients had lymphocytopenia with ALC <1,000/µL. All 6 patients treated in Cohort 1 had severe COVID-19 with Chest X-ray/CT-documented pneumonia associated with hypoxemia and respiratory distress, elevated baseline levels of inflammation markers in blood, such as abnormally high levels of CRP, IL-6, Ferritin, and/or LDH, and had a baseline score of 4 based on the 8-point ordinal scale (Table S2). By comparison, all 7 patients treated in Cohort 2 had critical COVID-19 with Chest X-ray/CT-documented pneumonia associated with hypoxemic respiratory failure requiring high flow oxygen therapy and/or non-invasive positive pressure ventilation (NIPPV), increased inflammation markers and had a baseline score of 3 based on the 8-point ordinal scale (Table S2). One Cohort 2 patient withdrew consent on day 4 and was replaced with a new patient to have a total of 6 patients evaluable for response.

### Ethics Statement and Study Approval

The study was performed according to the guidelines of the International Conference on Harmonization (ICH) for Good Clinical Practice (ICHE6/GCP). A written informed consent was obtained from patients prior to enrollment. The study protocol was approved by the WCG-Central Institutional Review Board (IRB) (OHRP/FDA Parent Organization number: IORG0000432; OHRP/FDA IRB registration number: IRB00000533). The Central IRB-approved study/protocol number was RPI015 (IRB Tracking Number: 20203418).

### Statistical Analyses of Clinical Data

Standard statistical methods were applied. Descriptive statics for patient cohorts were calculated using the R platform (R version 4.1.2 (2021-11-01)) ran in Rstudio (2021.09.0 Build 351) using standard functions provided by the statistical package “plyr_1.8.6”. Swimmer plots to visualize individual patient treatment outcomes and waterfall plots to visualize grouped and ranked outcomes were constructed using graph drawing packages implemented in the R programming environment: swimplot_1.2.0, colorspace_2.0-2, ggplot2_3.3.5, ggbreak_0.0.7, cowplot_1.1.1, ggh4x_0.2.1 and ggthemes_4.2.4. The Kaplan–Meier method was used to investigate the survival of patients utilizing software packages survival_3.2-13, survminer_0.4.9 and survMisc_0.5.5 in R environment. The kinetics of the CRS-related serum biomarkers were investigated using first-order exponential “decay” functions of percentage of each biomarker. The combined results for each biomarker were graphed on semi-log axis (log_10_ (% inhibition) = slope x Time + Intercept). The time to 50% reduction of the serum concentration of each biomarker was calculated from the intercept and the slope of the decay function ((log_10_ (50) – Intercept)/slope). These operations were carried out in the R programming environment: plyr_1.8.6 and ggplot2_3.3.5.

### LPS-GalN model of fatal cytokine storm and sepsis

The therapeutic activities of RJX, DEX, and RJX plus DEX were compared side by side in the LPS-GalN model of fatal CRS, sepsis and ARDS using male BALB/c mice, as previously described [26]. The RJX doses were 0.7 mL/kg (∼14 µL/20 g mouse) and 1.4 mL/kg (∼28 µL/20 g/mouse), and they were administered as intraperitoneal bolus injections. The doses of the active ingredients at the 0.7 mL/kg dose were 31.5 mg/kg Vitamin C, 2.2 mg/kg Vitamin B1, 0.09 mg/kg Vitamin B2, 4.2 mg/kg Vitamin B3, 0.1 mg/kg Vitamin B5, 4.2 mg/kg Vitamin B6, 0.07 mg/kg Vitamin B12, and 28.3 mg/kg magnesium sulfate. The doses for the active ingredients at the 1.4 mL/kg dose were 63 mg/kg Vitamin C, 4.4 mg/kg Vitamin B1, 0.18 mg/kg Vitamin B2, 8.4 mg/kg Vitamin B3, 0.2 mg/kg Vitamin B5, 8.4 mg/kg Vitamin B6, 0.14 mg/kg Vitamin B12, and 56.6 mg/kg magnesium sulfate [26]. These sub-MTD dose levels were based on the observation of single-agent activity in the LPS-GalN mouse model of sepsis [26]. 0.7 mL/kg mouse dose corresponds to a human equivalent dose (HED) of 0.057 ml/kg of RJX, which is 7.5% of its MTD of 0.759 mL/kg determined in the previously published randomized Phase 1 clinical trial in healthy volunteers [26]. The 1.4 mL per kg mouse dose corresponds to a human equivalent dose (HED) of 0.114 ml/kg of RJX, which is 15% of its MTD determined in the previously published randomized Phase 1 clinical trial in healthy volunteers.

The care and treatments of the animals were provided in accordance with the Guide for the Care and Use of Laboratory Animals. The study was approved by the Animal Care and Use Committee of Firat University (Number: 420629). Male BALB/c mice (6-8 weeks old and weighing on average 20 g), which were purchased from Laboratory Animal Center of Firat University, were randomly divided into different treatment groups, as previously reported [26]. Mice were housed in cages throughout the study, and the following conditions were maintained: temperature, 22 ± 2 °C; relative humidity, 55 ± 5%; and a 12-h light: dark cycle. As in previous studies, we applied the concealment of treatment allocation and blind outcome assessment to reduce the risk of bias [26]. In order to induce fatal CRS and ARDS, mice were challenged with an i.p. injection of LPS plus D-galactosamine (Sigma, St. Louis, MO). Each mouse received a 500 µL i.p. injection of LPS-GalN (consisting of 100 ng of LPS plus 8 mg of D-galactosamine). Treatments were delayed until 2 hours post-LPS-GalN injection when mice have a fulminant systemic inflammation with very severe lung and liver damage as well as markedly elevated inflammatory cytokine levels. Vehicle control mice were treated with 0.5 mL NS instead of RJX. NS was administered i.p 2 hours after LPS-GalN. Test mice received either RJX (0.7 ml/kg or 1.4 ml/kg) or DEX (0.1 mg/kg, 0.6 mg/kg or 6.0mg/kg) as a monotherapy in a side-by-side comparison. We also examined a combination of 0.7 mL/kg RJX with either 0.6 mg/kg or 6 mg/kg DEX at 2 hours post-LPS-GalN injection. Drugs were administered i.p. in a total volume of 0.5 ml. The human equivalent dose (HED) levels were determined as described [37]. Tissue levels of oxidative stress biomarkers superoxide dismutase (SOD), catalase (CAT) and glutathione peroxidase (GSH-Px), ascorbic acid levels, and malondialdehyde (MDA) were determined to examine the effects of RJX, DEX, and RJX + DEX on the sepsis-associated oxidative stress. Levels of Interleukin 6 (IL-6) and tumor necrosis factor-alpha (TNF-α) in serum samples were measured by quantitative ELISA using the commercially available Quantikine ELISA kits, as previously reported [26]. At the time of death, blood samples were obtained for biomarker studies. In addition, lungs and liver were harvested, fixed in 10% buffered formalin, and processed for histopathologic examination. 3 μm sections were cut, deparaffinized, dehydrated, and stained with hematoxylin and eosin (H & E) and examined with light microscopy using a digital image capture camera by an experienced histopathologist who was blind to the study groups.

### Statistical analysis of Non-Clinical Data

Statistical analyses were performed using standard methods and the SPSS statistical program (IBM, SPPS Version 21), as previously reported [26]. P-values <0.05 were considered significant.

## Results

### Non-clinical Proof of Concept Data Supporting the Use of RJX Alone and In Combination with DEX for Treatment of Severe COVID-19

LPS-GalN-challenged mice experience a rapid onset systemic inflammation with markedly elevated inflammatory cytokine levels as well as severe lung injury, reminiscent of severe COVID-19, as early as 2 hours after the administration of LPS-GalN [26]. A single low dose RJX at the assigned low dose levels of 0.7 ml/kg (HED: 0.057 ml/kg; 7.5% of human MTD) delivering 31.5 mg/kg Vitamin C, 2.2 mg/kg Vitamin B1, 0.09 mg/kg Vitamin B2, 4.2 mg/kg Vitamin B3, 0.1 mg/kg Vitamin B5, 4.2 mg/kg Vitamin B6, 0.07 mg/kg Vitamin B12, and 28.3 mg/kg magnesium sulfate) and 1.4 ml/kg (HED: 0.114 ml/kg; 15% of human MTD) delivering 63 mg/kg Vitamin C, 4.4 mg/kg Vitamin B1, 0.18 mg/kg Vitamin B2, 8.4 mg/kg Vitamin B3, 0.2 mg/kg Vitamin B5, 8.4 mg/kg Vitamin B6, 0.14 mg/kg Vitamin B12, and 56.6 mg/kg magnesium sulfate, which was administered at 2 h after LPS-GalN injection, effectively reversed the LPS-GalN induced increased serum levels of the pro-inflammatory cytokines IL-6 and TNF-α within 24 h (Supplemental Information, Figure S2). 1.4 ml/kg low dose (15% of MTD) RJX was significantly more effective than a single DEX injection at standard 0.6 mg/kg dose level (HED: 0.05 mg/kg; 4 mg standard dose for an 80 kg person) and comparable to 6.0 mg/kg high dose DEX (HED: 0.49 mg/kg; 39 mg dose for an 80 kg person, which is higher than the standard 4-8 mg dose levels for DEX) (Figure S2). The observed reversal of the inflammatory cytokine response by RJX (0.7 or 1.4 mL/kg) or DEX (0.6 or 6.0 mg/kg) was associated with a partial reversal of the lung and liver injury that was documented at 2 h post-LPS-GalN injection when treatments were initiated (Supplemental Information, Figure S3, Figure S4, Figure S5) as well as a significant improvement of the survival outcome in this LPS-GalN model of otherwise invariably fatal CRS and ARDS (Supplemental Information, Figure S6). A combination of low dose (0.7 ml/kg) RJX and high dose (6.0 mg/kg) DEX effectively reversed the increased serum levels of the systemic inflammation markers (IL-6, TNF-α, and LDH) within 24 h, and appeared to be overall more effective than DEX alone or RJX alone (Figure S7).

LPS-GalN caused severe inflammation and oxidative stress in the lungs and liver with enhanced lipid peroxidation, as measured by a marked elevation of MDA levels, and decreased tissue levels of Vitamin C as well as the anti-oxidant enzymes GSH-Px that inhibits lipid peroxidation and SOD that converts superoxide anion radicals contributing to lipid peroxidation into hydrogen peroxide and oxygen. Low dose RJX alone or in combination with DEX significantly suppressed the oxidative stress, as documented by increases in Vitamin C, GSH-Px and SOD levels that were markedly reduced by LPS-GalN (Figure S8). The tissue healing activity of the combination therapy in the lungs and liver was also more pronounced than the tissue healing activity of RJX alone or DEX alone (Figure S9, Figure S10). In contrast to the rapid death of all control mice treated with vehicle/normal saline (NS) (Median survival: 4.3 h post-LPS-GalN or

2.3 h after administration of NS), 100% of mice treated with low dose RJX + high dose DEX survived the LPS-GalN challenge (Median survival: >24 hours post LPS-GalN or >22 hours after initial administration of RJX+DEX) (Figure S11). By comparison, the combined group of mice treated with monotherapy (i.e., RJX alone or DEX alone) (n=12) had a 24-hour survival rate of 41.7% (Monotherapy with RJX or DEX vs. Combination therapy with RJX+DEX: Log-rank X^2^ = 3.053, p=0.081).

We next evaluated the efficacy of a combination regimen that employs low dose RJX (0.7 mL/kg) plus standard dose (0.6 mg/kg) DEX. Notably, delayed treatments with low dose RJX plus standard dose DEX starting at 2 h after the LPS-GalN injection reversed the lung damage, as evidenced by significantly reduced histopathological lung scores (Figure 1). The tissue healing activity of the combination was more pronounced than the tissue healing activity of RJX alone or DEX alone (Figure 1A). Hence, although treatments were delayed until the onset of fulminant cytokine storm and systemic inflammation as well as very severe lung damage, a near-complete recovery of the inflammatory lung injury was achieved in the vast majority of mice within 24 hours. Similar to its effects on the LPS-GalN induced ALI, RJX + DEX combination significantly reduced the liver injury (Figure 1B). While 5 of 10 mice (50 %) treated with standard dose DEX alone and 14 of 20 mice (70%) treated with low dose RJX alone had severe residual lung injury (histopathological lung score ≥3), only 2 of 20 mice (10%) of mice treated with the DEX+RJX combination had severe residual lung damage. The observed superiority of the combination regimen was statistically significant (Figure 1C). Similarly, while 5 of 10 DEX treated mice (50%) and 12 of 20 RJX treated mice (60%) had severe residual liver damage, only 1 of 20 mice (10%) treated with the DEX+RJX combination had severe residual liver damage and this difference was statistically significant (Figure 1C). In contrast to the rapid death of all control mice treated with NS (Median survival: 5.1 h post LPS-GalN or 3.1 h after administration of NS), 15 of 20 (75%) of mice treated with RJX + DEX survived the LPS-GalN challenge (Median survival: >24 h post LPS-GalN (Figure 2). By comparison, the combined group of mice treated with monotherapy (i.e., RJX alone or DEX alone) (N=30) had a 24-h survival rate of 53% (i.e., 16 of 30 mice) (Monotherapy with RJX or DEX vs. Combination therapy with RJX+DEX: Log-rank X^2^ = 3.977, p=0.046).

**Figure 1.**
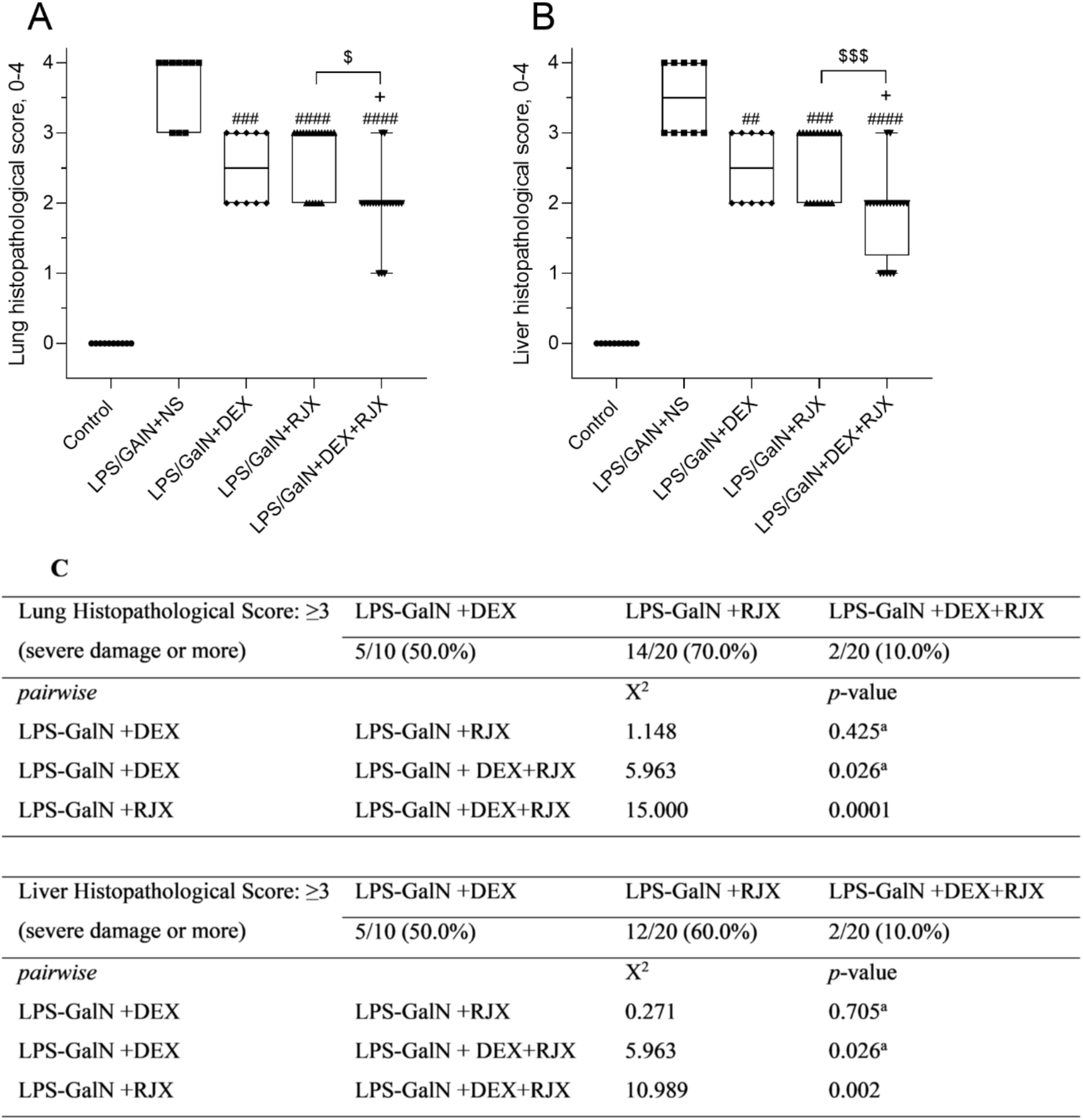
In Vivo Treatment Activity of Rejuveinix (RJX), Dexamethasone (DEX), and RJX+DEX on Lung and Liver Histopathological Scores in the LPS-GalN Mouse Model of Fatal Cytokine Storm, Sepsis, Systemic Inflammation, ARDS and Multiorgan Failure. BALB/C mice were treated with i.p injections of RJX (n=20, 6-fold diluted, 4.2 mL/kg, 0.5 ml/mouse), DEX (n= 10, 6 mg/kg, 0.5 mL/mouse), RJX + DEX (n=20, 0.5 mL/mouse), or vehicle (NS, 0.5 mL/mouse) two hours post-injection of LPS-GalN. Except for untreated control mice (Control, n=10), each mouse received 0.5 ml of LPS-GalN (consisting of 100 ng of LPS plus 8 mg of D-galactosamine) i.p. The depicted Whisker plots represent the median and values. In (A), the lung histopathological score (“lung injury score”) was graded according to a 5-point scale from 0 to 4 as follows: 0, l, 2, 3, and 4 represented no damage, mild damage, moderate damage, severe damage, and very severe damage, respectively. In (B), the liver histopathological score (“liver injury score”) was graded according to a 5-point scale from 0 to 4 as follows: 0, l, 2, 3, and 4 represented no damage, mild damage, moderate damage, severe damage, and very severe damage, respectively. Kruskal-Wallis test and Mann Whitney U test were used for comparing the results among different treatment groups. Statistical significance between groups is shown by ## p<0.01; ### p<0.001; #### p<0.0001 as compared to LPS/GaIN+NS group, + p<0.05 as compared to LPS/GaIN+DEX group, and $ p<0.05; $$$ p<0.001 pairwise comparisons between the groups. In (C), for severe damage, lung and liver histological scores were compared by Pearson’s Chi-Square or Fisher’s Exact test. ^a^ Fisher’s Exact Chi-Square test used.

**Figure 2.**
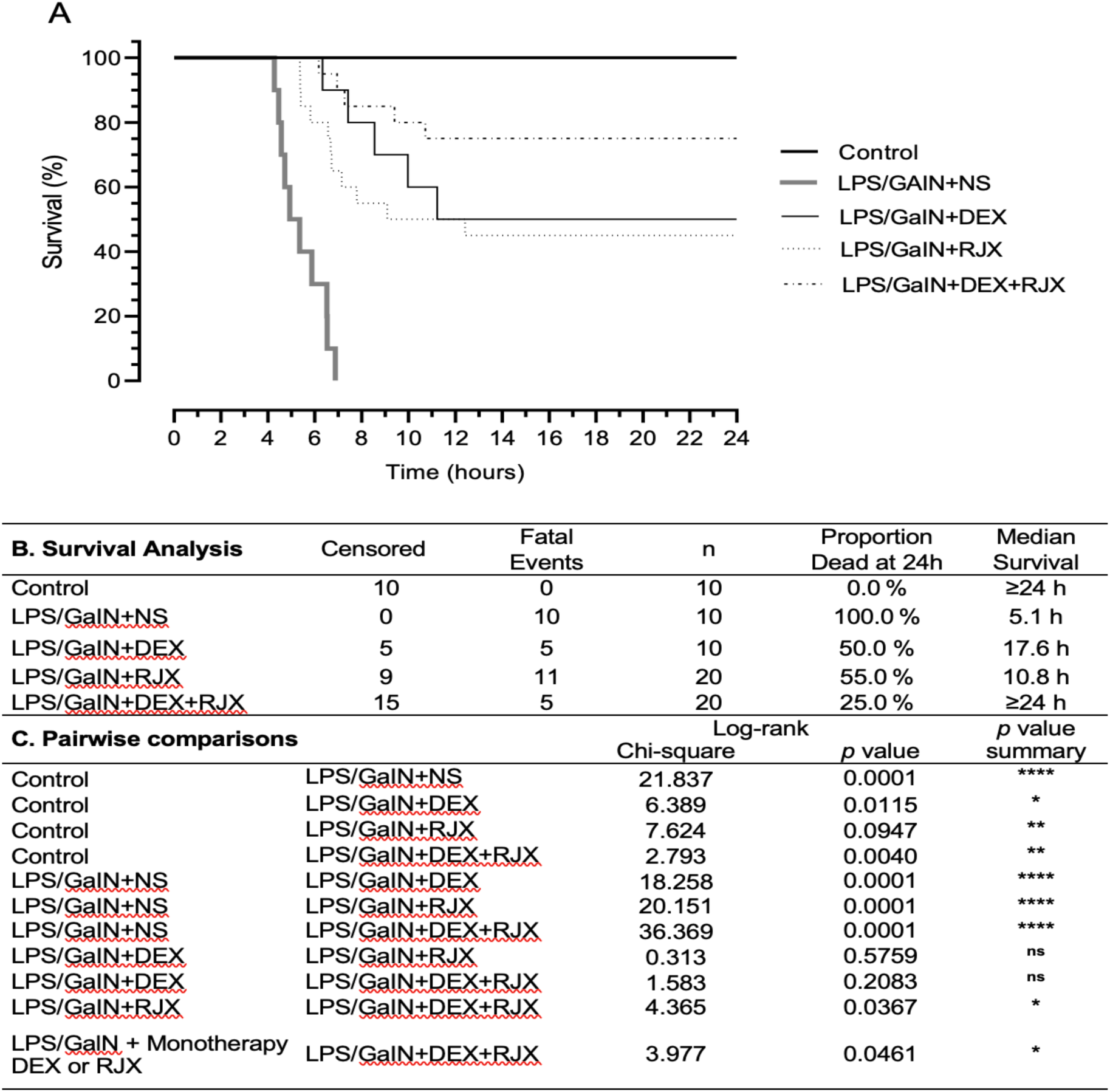
In Vivo Treatment Activity of Low Dose Rejuveinix (RJX), Standard Dose Dexamethasone (DEX), and Their Combination in the LPS-GalN Mouse Model of Fatal Cytokine Storm, Sepsis, Systemic Inflammation, ARDS and Multiorgan Failure. BALB/C mice were treated with i.p injections of RJX (n=20, 6-fold diluted, 4.2 mL/kg, 0.5 ml/mouse), DEX (n= 10, 0.6 mg/kg, 0.5 mL/mouse), RJX + DEX (n=20, 0.5 mL/mouse), or vehicle (NS, 0.5 mL/mouse) two hours post-injection of LPS-GalN. Except for untreated control mice (Control, n=10), each mouse received 0.5 ml of LPS-GalN (consisting of 100 ng of LPS plus 8 mg of D-galactosamine) i.p. The cumulative proportion of mice remaining alive (Survival, %) is shown as a function of time after the LPS-GalN challenge. Depicted are the Kaplan Meier survival curves (Panel A) and survival data with statistical analysis (Panels B and C) of the different treatment groups.

### Clinical Proof of Concept Data Supporting the Use of RJX in Combination with SOC for Treatment of Severe COVID-19

#### Safety

All patients were enrolled ≥3 months prior to the identification of the first confirmed U.S case of the Omicron variant. Patient characteristics and demographic features are shown in Table S2. All patients were treated daily with IV RJX plus institutional SOC (Table S3). One patient withdrew consent on day 4 without any treatment-emergent AEs. Table S4 details all Grade 3-5 AEs reported. No patient developed a DLT or SAE while still receiving RJX. None of the reported Grade 3-5 AEs was deemed related to RJX – they were all reported as related to COVID-19. Listing of all grade SAEs is provided in Table S5. Table S6 depicts the incidence of Grade 3-5 AEs and SAEs by MedDRA PT and listing of deaths for all enrolled patients.

#### Pharmacodynamics

Protocol therapy resulted in a rapid resolution of pneumonia and systemic inflammation in 9 of 12 patients enrolled. In these 9 patients, we observed a rapid normalization in the levels of the inflammation biomarkers CRP, Ferritin, LDH, and IL-6. We investigated the first order kinetics of the CRP decline in each of the 9 responding patients by fitting a straight line to a semi-log plot of the portion of the biomarker curve over the course of RJX treatment when a maximum reduction in values was observed. The slope of the line represents the rate constant for CRP reduction in log_10_ scale, and times to 50% reduction of CRP values were calculated using the rate constant (Figure 3). Serum CRP levels showed a rapid decline with an average (Mean±SE) time of 1.6±0.2 days to reach 50% of baseline values (Median: 1.7 days, Range: 0.9-2.5 days) (Figure S12). Similar results were observed for IL-6 (N=5; Mean: 13.1 ± 6.1 days, Median: 6.3 days, Range: 1.6 - 29.3 days (Figure 4), Ferritin (N=8; Mean: 8.8 ± 3.4 days, Median: 5.2 days, Range: 2.9 - 32.1 days) (Figure S13) and LDH (N=5; Mean: 8.6 ± 1.8 days, Median: 7.2 days, Range: 4.9 - 14.6 days) levels (Figure S14). By comparison, the kinetics of normalization of elevated TGF-β levels was much slower and near-normal levels were achieved in some patients only after 1-2 months (N=4; Mean±SE: 26.6±17.3 days; Median 13.2 days; Range: 2.3-77.6 days) (Figure S15).

**Figure 3.**
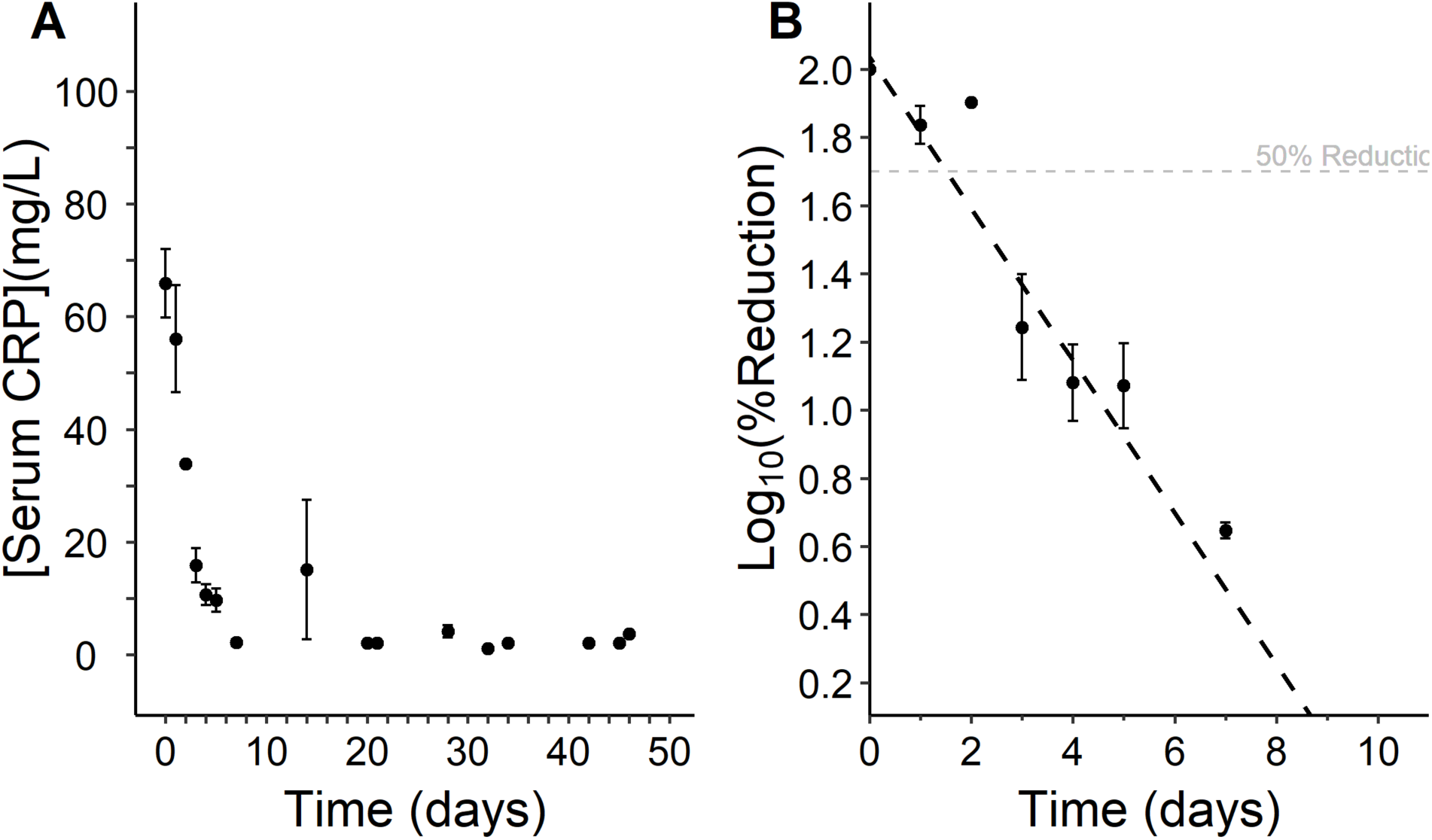
Time-dependent decrease of CRP Values in RJX-Treated Severe COVID-19 Patients. [A] Serum CRP concentration is depicted over 46 days for 9 evaluable responders [B] The first order kinetics of the reductions in serum CRP values was investigated by fitting a straight line to a semi-log plot of the portion of the CRP concentration x time curve that displayed maximum reduction in CRP values over the course of RJX treatment. There were 36 independent data points across 7 days. The slope of the line represents the rate constant for CRP reduction in log_10_ scale (viz.: -0.2231). Time to 50% reduction (T_50_ = 1.5 days) of serum CRP values was calculated using the rate constant and intercept value of 2.04. Equation of the line: Log_10_ (% Reduction) = -0.22309 x Time(Days) + 2.04.

**Figure 4.**
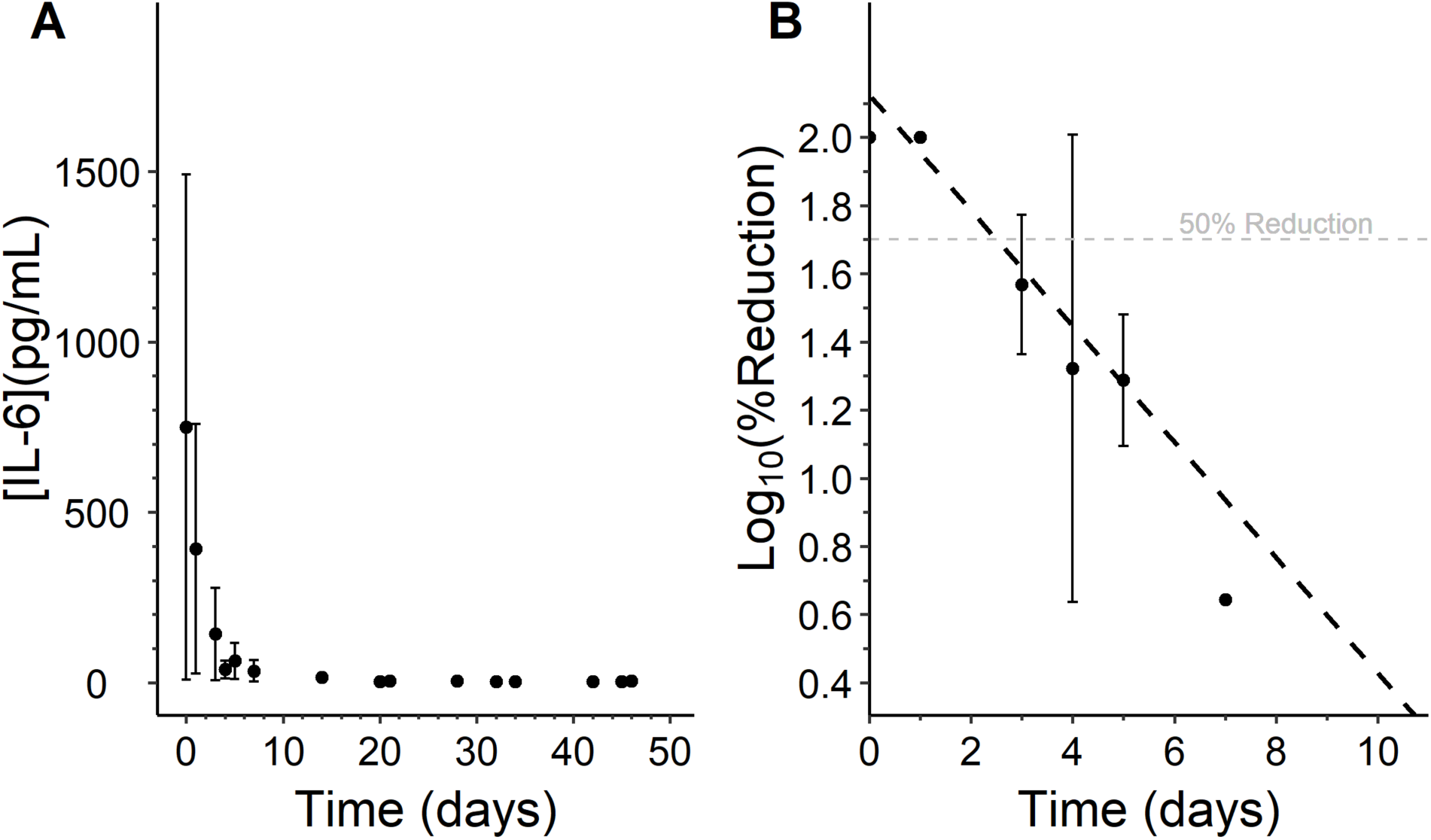
Time-dependent Reduction of Serum IL-6 Concentration in RJX-Treated Severe COVID-19 Patients. [A] Serum IL-6 concentration is depicted over 46 days for 5 evaluable responders [B] The first order kinetics of the reductions in serum IL-6 values was investigated by fitting a straight line to a semi-log plot of the portion of the IL-6 concentration x time curve that displayed maximum reduction in IL-6 values over the course of RJX treatment. There were 18 independent data points across 7 days. The slope of the line represents the rate constant for IL-6 reduction in log_10_ scale (viz.: -0.1698). The time to 50% reduction (T_50_ = 2.5 days) of serum IL-6 values was calculated using the rate constant and the intercept value of 2.13. Equation of the line: Log_10_ (% Reduction) = -0.16983 x Time(Days) + 2.13.

#### Efficacy

The outcome data for 12 evaluable patients are shown in Table 1 (6 from Cohort 1 and 6 from Cohort 2; Patient 008-1204 who withdrew his consent on day 4 is not shown in this table). Nine (9) of the 12 patients, including each of the 6 Cohort 1 patients and 3 of 6 Cohort 2 patients with hypoxemic respiratory failure, showed rapid clinical recovery with normalization of the blood oxygen levels. The overall survival curve is shown in Figure 5; the median survival was not reached. Figure 6 shows the kinetics of improvement or worsening of the clinical health status scores according to the 8-point ordinal scale for each patient. The score changes were charted utilizing Swimmer plots. Figure S16 shows a waterfall plot depicting the maximum increase (improvement) or decrease (worsening) of the 8-point Ordinal Scale scores. The score change ranged from -2 to +5 (Median: +3.5; Mean ± SE: 2.4 ± 0.8). The score change for Cohort 1 patients (N=6) ranged from +3 to +4 (Median: +3.5, Mean ± SE : 3.5± 0.2). By comparison, the score change for Cohort 2 patients (N=6) ranged from -2 to +5 (Median: +1; Mean ± SE: 1.3±1.5). Each of the 6 patients in Cohort 1 had a baseline score of 4. Their oxygen supplementation requirements have resolved, and they were consequently discharged from the hospital between 3 to 7 days (median: 4.5 days) after initiation of protocol therapy. Hence, their scores showed a +3 (Score 7: Not hospitalized, limitation on activities and/or requiring home oxygen) to +4 improvement within a week (Score 8: Not hospitalized, no limitations on activities – corresponds to ECOG score of 0). Of the 6 patients with hypoxemic respiratory failure who had a baseline score of 3 and required high flow oxygen and/or NIPPV, 3 were discharged from hospital at 7,7, and 14 days respectively upon resolution of their hypoxemic respiratory failure with a +4-+5 improvement on the 8-point ordinal scale (Figure 6). Two of the remaining 3 patients experienced progression of their COVID-19 requiring intubation on day 8 and one patient developed COVID-19 related complications, including sepsis with coagulopathy (day 10-17), pulmonary embolism (day 19), and fatal mesenteric ischemia (day 23) (Figure 6, Table S6).

**Table 1:** Treatment Outcome of Patients in Part 1 of RPI015 Study.

| Patient No. | Cohort# | Progression of COVID-19 and NIV Failure | Required IMV | Time to IMV (Time to death) | Clinical Improvement |  | CXR/CT Improvement at Discharge from Hospital | Recovery on 8-point Ordinal Scale | Patient status at the last follow-up date |  |  |
| --- | --- | --- | --- | --- | --- | --- | --- | --- | --- | --- | --- |
|  |  |  |  |  | Time from 1<sup>st</sup> dose of RJX to Resolution of HRF (Time to Discharge) in Days | COVID-19 Symptoms Improved/Resolved |  |  | Last follow-up date (Days from ICF) | Status (Alive/Dead) | ECOG Performance Status |
| 002-1102 | 1 | No | No | NA(NA) | NA (4) | +/+ | NA | +3 | 65 | Alive | 0 |
| 002-1203 | 2 | Yes | Yes | 8 (26) | NA (NA) | -/- | NO | -2 (no recovery) | 26 | Dead | 5 |
| 002-1104 | 1 | No | No | NA(NA) | NA (3) | +/+ | NA | +3 | 64 | Alive | 0 |
| 008-1201 | 2 | No | No | NA(NA) | 5 (7) | +/+ | YES | +5 | 68 | Alive | 0 |
| 008-1103 | 1 | No | No | NA(NA) | NA (5) | +/+ | YES | +3 | 65 | Alive | 0 |
| 008-1105 | 1 | No | No | NA(NA) | NA (5) | +/+ | NO | +4 | 65 | Alive | 0 |
| 008-1208 | 2 | No | No | NA (NA) | 3 (7) | +/+ | YES | +5 | 66 | Alive | 0 |
| 008-1210 | 2 | Yes | Yes | 8 (14) | NA (NA) | -/- | NA | -2 (no recovery) | 14 | Dead | 5 |
| 007-1201 | 2 | No* | No | 21 (21) | NA (NA) | -/- | NA | -2 (no recovery) | 21 | Dead | 5 |
| 007-1202 | 2 | No | No | NA (NA) | 14 (14) | +/-<sup>a</sup> | NA | +4 | 60 | Alive | 0 |
| 008-1111 | 1 | No | No | NA (NA) | NA (3) | +/+ | NO | +3 | 65 | Alive | 0 |
| 007-1103 | 1 | No | No | NA (NA) | NA (7) | +/+ | NA | +4 | 60 | Alive | 0 |

Covid-19: Corona virus disease 2019; NIV: Non-invasive ventilation; IMV: Invasive mechanical ventilation; ICF: Informed consent form; RJX: Rejuveinix; <sup>a</sup>007-1202 reported mild residual shortness of breath at last follow-up
8-point Ordinal Scale:
1. Death 2. Hospitalized, on invasive mechanical ventilation or ECMO 3. Hospitalized, on non-invasive ventilation or high flow oxygen devices 4. Hospitalized, requiring supplemental oxygen 5. Hospitalized, not requiring supplemental oxygen - requiring ongoing medical care (COVID-19 related or otherwise) 6. Hospitalized, not requiring supplemental oxygen - no longer requires ongoing medical care 7. Not hospitalized, limitation on activities and/or requiring home oxygen 8. Not hospitalized, no limitations on activities
ECOG Activity Level:

**Figure 5.**
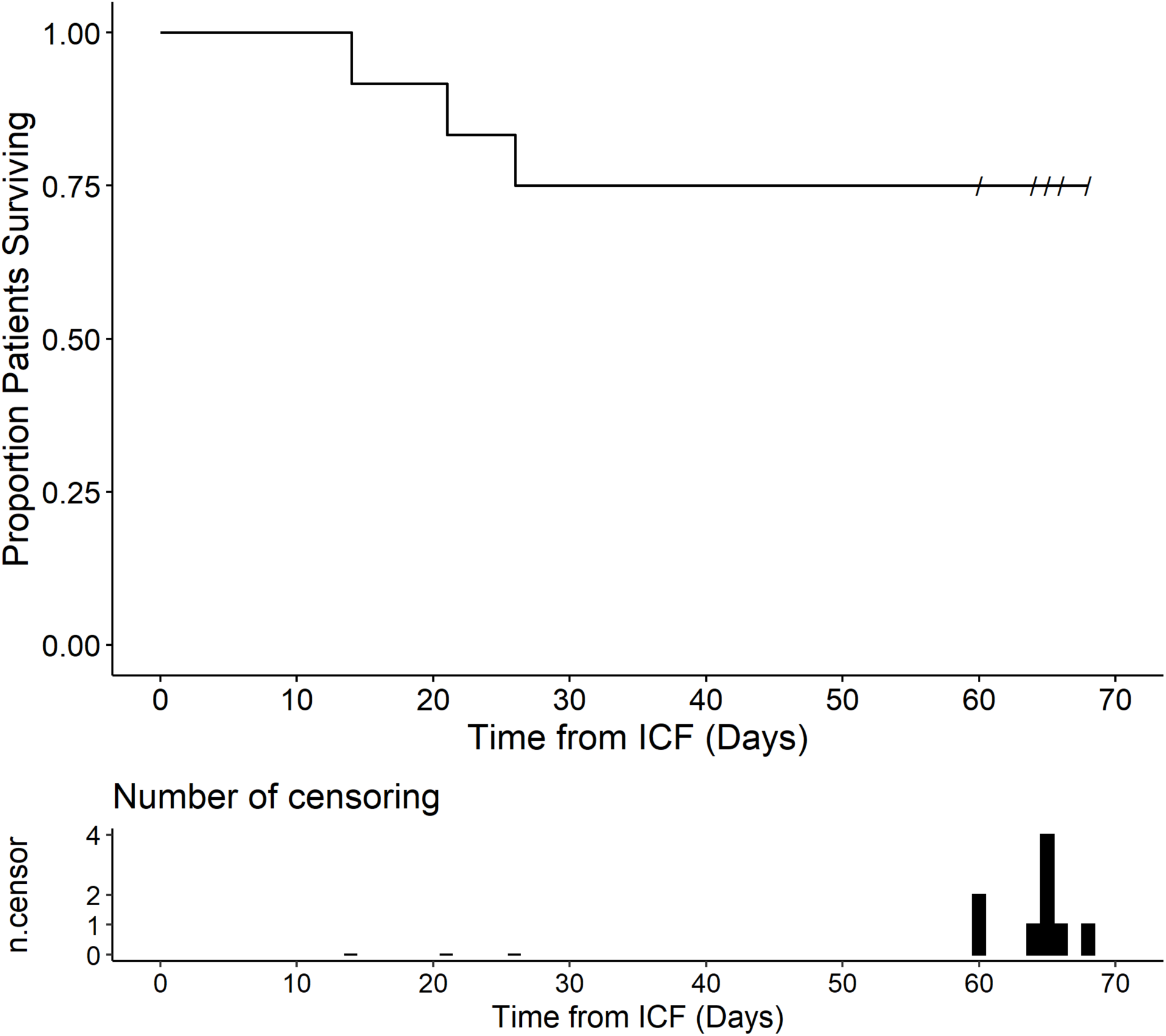
Survival Outcome of Severe COVID-19 Patients Treated with RJX Plus SOC. Depicted is the overall survival curve of the 12 hospitalized COVID-19 patients shown in Figure 3 along with the corresponding censor plot. All patients were treated with RJX + institutional SOC. Nine patients recovered and remained alive for ≥60 days.

**Figure 6.**
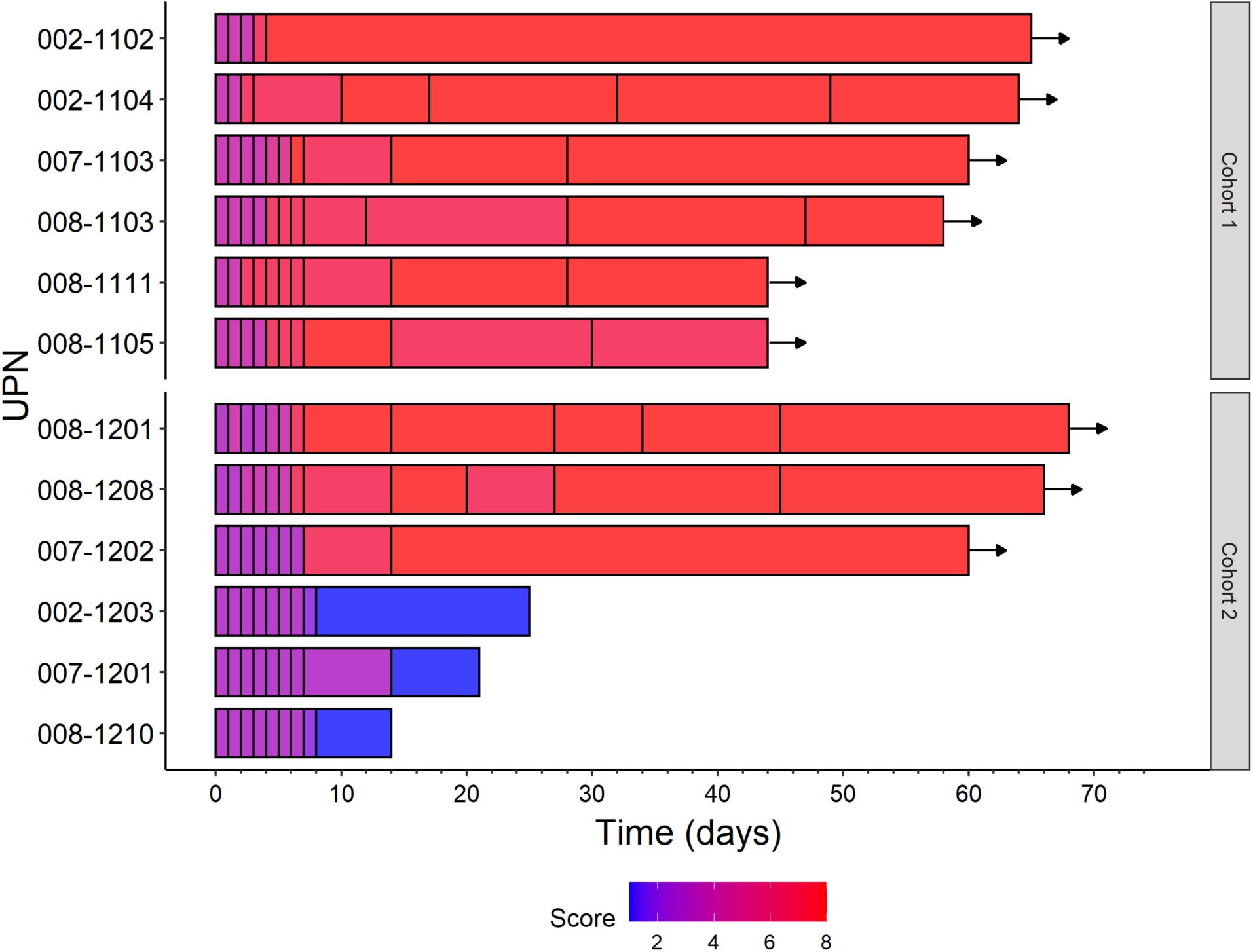
Swimmer plot of clinical health score changes in RJX treated COVID-19 patients. Scores according to the 8-point ordinal scale are indicated with different colors. Arrow: Alive.

## Discussion

Globally, COVID-19 now is a major cause of mortality [9, 37-41]. Patients with high-risk COVID-19 with viral sepsis are in urgent need of effective strategies that can prevent and/or reverse the systemic inflammatory process and its complications, including acute respiratory distress syndrome (ARDS) and multi-organ failure [42-53]. Here we report a pilot clinical study of RJX plus standard of care in hospitalized COVID-19 patients with pneumonia ≥3 months prior to the identification of the first confirmed U.S case of the Omicron COVID-19 variant, which extends the preclinical observations in a mouse model of fatal ARDS and provides promising early clinical proof of concept data for developing RJX as part of the multi-modality treatment strategies against severe COVID-19.

DEX has been shown to improve the survival outcome of patients with ARDS [53-60]. Furthermore, recent clinical studies, including the open-label randomized “RECOVERY” trial, have demonstrated that the standard anti-inflammatory drug DEX improves the clinical course and treatment outcome of hospitalized high-risk COVID-19 patients [54-57] Rees et al. reported that DEX reduced the death rate by one third in patients on a ventilator and by one fifth in patients on oxygen support [55]. Early findings showed that DEX decreased the mortality risk from 40% to 28% in ventilated patients and 25%–20% for COVID-19 patients on oxygen therapy over 28 days [57]. COVID-19 patients with moderate or severe ARDS who were treated with DEX plus SOC had significantly more ventilator-free days over 28 days than the reference group treated with SOC alone [58].

IL-1, IL-6, TNF-α, and TGF-β are important pro-inflammatory cytokines that contribute to the pathophysiology of the cytokine release syndrome (CRS), ARDS and multi-organ failure in critically sick adult COVID-19 patients with viral sepsis [13-23] as well as children and adolescents with COVID-19 who develop a multi-system inflammatory syndrome (MIS-C) [59-62]. Recently, we demonstrated that RJX prevents in the LPS-GalN mouse model of sepsis the marked increase of each of these cytokines in the serum as well as lungs and liver [26]. Furthermore, preliminary evidence suggested that it may also be effective in reversing sepsis if administered after the onset of sepsis [35].

The primary objective of the non-clinical proof-of-concept study was to compare the effects of RJX, DEX, and their combination on the severity of CRS and survival outcome in an animal model that employs LPS-GalN to induce fatal sepsis, CRS, ARDS. We hypothesized that RJX, especially when combined with DEX, would improve the survival outcome of mice who develop systemic inflammation following injection with a lethal dose of LPS-GalN. The experimental data demonstrate that RJX – at a dose level >10-times lower than its clinical MTD - exhibits potent single-agent anti-inflammatory activity and is as effective as DEX at standard dose levels in this model of fatal sepsis. The combination of RJX plus DEX immediately and profoundly decreased the inflammatory cytokine responses to LPS-GalN, mitigated the inflammatory tissue damage in the lungs and liver, and prevented a fatal outcome. Although the treatments were started after the onset of fulminant cytokine storm characterized by markedly elevated serum cytokine levels and systemic inflammation as well as severe lung damage, a near-complete recovery of the inflammatory lung injury was achieved within 24 hours, and the survival outcome was improved. The superiority of the combination regimen was particularly striking when we compared the histopathological data on sepsis-associated organ damage in lungs and liver from mice treated with monotherapy (RJX or DEX alone) vs. combination therapy. Notably, initiation of combination therapy after onset of the inflammatory cytokine responses and systemic inflammation resulted within 24 hours in a near-complete recovery of very severe organ damage in the lungs that was caused by LPS-GalN induced sepsis. These findings provide the preclinical proof of concept for the clinical development of RJX as an adjunct of the standard of care in the multi-modality management of sepsis and its complications.

Several monoclonal antibodies that serve as inhibitors of the receptors and/or signaling pathways activated by specific pro-inflammatory cytokines are being developed and evaluated as anti-inflammatory drug candidates for prevention of CRS-related complications of severe COVID-19, including but not limited to the IL-1 receptor antagonist Anakinra (Kineret), IL-6 receptor antagonist tocilizumab (Actemra), TNF-α inhibitor infliximab (Remicade) [14,15,22,42,49-52]. A major potential advantage of RJX versus biotherapeutic agents such as monoclonal antibodies that inhibit specific cytokine pathways is its ability to inhibit multiple pathways combined with its low cost of production which should make it readily available in low-to middle-income countries and therefore its further development and assessment as a potential COVID-19 drug candidate has potential global relevance.

RJX is hoped to reduce the case mortality rate of severe COVID-19 by accelerating the resolution of the systemic and pulmonary inflammatory process. Here we showed that 9 of the 12 patients, including 3 patients with hypoxemic respiratory failure, showed rapid clinical recovery with resolution of the hyperinflammatory response and oxygen therapy requirements. This early clinical experience with RJX in 12 hospitalized patients with severe-critical COVID-19 warrants further investigation of its clinical impact potential. We hypothesize that the addition of RJX to a standard of care, including DEX, will shorten the time to clinical resolution of the cytokine-mediated multi-system inflammatory process and thereby mitigate the inflammatory organ injury. RJX has no steroids, and it contains niacinamide, pyridoxine, cyanocobalamin and Mg-sulfate in addition to ascorbic acid and thiamine. Additional studies in a randomized setting are needed to confirm the reported preliminary findings and assess any potential benefit from the addition of RJX to the SOC. RJX is currently being evaluated for its clinical impact potential as a component of multi-modality treatment algorithms for severe to critical COVID-19 in a placebo-controlled randomized Phase II study (ClinicalTrials.gov Identifier: NCT04708340). The randomized trial will test the hypothesis that RJX will contribute to a faster resolution of HRF when combined with DEX and other components of the standard of care.

## Data Availability

The original contributions presented in the study are included in the article/Supplementary Material. Further inquiries can be directed to the corresponding author.

## Permission to reproduce material from other sources

Not applicable

## Ethics Statement and Study Approval

The clinical study protocol was approved by the WCG-Central Institutional Review Board (IRB) (OHRP/FDA Parent Organization number: IORG0000432; OHRP/FDA IRB registration number: IRB00000533). The Central IRB-approved study/protocol number was RPI015 (IRB Tracking Number: 20203418). The study was performed in compliance with the International Conference on Harmonization (ICH) guidelines for Good Clinical Practice (ICHE6/GCP). Each patient provided a written informed consent (ICF) prior to enrollment.

The non-clinical research project was approved by the Animal Care and Use Committee of Firat University (Project No. 03092020-391-047; Ethics Committee Number: 420629).

## Author Contributions

Each author has made significant and substantive contributions to the study, reviewed and revised the manuscript, provided final approval for submission of the final version. F.M.U conceived the study, designed the evaluations reported in this paper, directed the data compilation and analysis, analyzed the data, and prepared the initial draft of the manuscript. K.S. directed the non-clinical study. K.S. and C.O. collected the non-clinical data and performed their analysis. I.H.O. performed the necropsies and histopathologic examinations on mice. M.A., A.S., A.W.S, J.V assisted in clinical study design, served as site investigators, screened and recruited participants, administered treatments, assessed adverse events and disease responses, collected safety and efficacy data, met regularly to review study data during the study, and reviewed the manuscript; F.M.U., S.Q., and C.L. analyzed and validated clinical data, performed statistical analyses.

## Acknowledgments

The authors thank the patients who participated in this trial and their families, the coinvestigators, nurses, and study coordinators at each of the participating sites. The study was performed at the following centers in the US as an open-label study sponsored by Reven Pharmaceuticals: (1) Memorial Hermann Memorial City Medical Center, Houston, Texas; (2) Christus Health Santa Rosa Hospital, New Braunfels, Texas; (3) United Memorial Medical Center, Houston, Texas; (4) PRX Research and Dallas Regional Medical Center, Dallas, Texas.

This study was sponsored by Reven Pharmaceuticals, which provided RJX. We thank the Reven team members for their contributions to clinical operations, quality assurance, and IP/vendor management.

