## Supplemental information for "Clinical and Non-clinical Proof of Concept Supporting the Development of RJX As an Adjunct to Standard of Care Against Severe COVID-19"

#### Supplemental Materials and Methods

##### Rejuveinix (RJX)

RJX is a pharmaceutical composition containing several anti-inflammatory and anti-oxidant vitamins, including Vitamin C, Vitamin B1, Vitamin B2, Vitamin B3, Vitamin B5, Vitamin B6, and Vitamin B12. RJX is being developed as an anti-inflammatory and anti-oxidant pharmaceutical composition for COVID-19 patients at high risk for viral sepsis and ARDS [17,18]. The RJX drug product is comprised of two vials (**Table S1**): Sodium bicarbonate-containing Vial B and Vial A containing the following active ingredients: Ascorbic acid USP (Vitamin C): 899.33 mg/10 mL Vial A = 449.7 mg/10 mL RJX after 1:1 dilution with Vial B), Thiamine HCl USP (Vitamin B1): 63.33 mg/10 mL Vial A = 31.7 mg/10 mL RJX after 1:1 dilution with Vial B), Riboflavin 5'phosphate USP (Vitamin B2): 2.53 mg/10 mL Vial A = 1.265 mg/10 mL RJX after 1:1 dilution with Vial B, Niacinamide USP (Vitamin B3): 118.80 mg/10 mL Vial A = 59.4 mg/10 mL RJX after 1:1 dilution with Vial B, calcium D-pantothenate USP (Vitamin B5): 2.93 mg/10 mL Vial A = 1.465 mg/10 mL RJX after 1:1 dilution with Vial B, Pyridoxine HCl USP (Vitamin B6) : 118.80 mg/10 mL Vial A = 59.4 mg/10 mL RJX after 1:1 dilution with Vial B, Cyanocobalamin Crystalline USP (Vitamin B12): 1.93 mg/10 mL Vial A 0.965 mg/10 mL RJX after 1:1 dilution with Vial B, and magnesium sulfate USP: 808.00 mg/10 mL Vial A = 404 mg/10 mL RJX after 1:1 dilution with Vial B [10]. In the clinical study, for both cohorts, 6 consecutive patients received a fixed dose of 20 mL RJX (10 mL of Vial A plus 10 mL of Vial B) mixed with 100 mL normal saline (total volume = 120 mL) which was administered by IV infusion over a period of 40 +/- 10 min daily.

In the non-clinical proof of concept study in mice, the RJX doses were 0.7 mL/kg (~14 μL/20 g mouse) and 1.4 mL/kg (~28 μL/20 g/mouse) and they were administered as intraperitoneal bolus injections. The doses of the active ingredients at the 0.7 mL/kg dose were 31.5 mg/kg Vitamin C, 2.2 mg/kg Vitamin B1, 0.09 mg/kg Vitamin B2, 4.2 mg/kg Vitamin B3, 0.1 mg/kg Vitamin B5, 4.2 mg/kg Vitamin B6, 0.07 mg/kg Vitamin B12, and 28.3 mg/kg magnesium sulfate. The doses for the active ingredients at the 1.4 mL/kg dose were 63 mg/kg Vitamin C, 4.4 mg/kg Vitamin B1, 0.18 mg/kg Vitamin B2, 8.4 mg/kg Vitamin B3, 0.2 mg/kg Vitamin B5, 8.4 mg/kg Vitamin B6, 0.14 mg/kg Vitamin B12, and 56.6 mg/kg magnesium sulfate [26]. These sub-MTD dose levels were based on the observation of single-agent activity in the LPS-GalN mouse model of sepsis [26]. 0.7 mL/kg mouse dose corresponds to a human equivalent dose (HED) of 0.057 ml/kg of RJX, which is 7.5% of its MTD of 0.759 mL/kg determined in the previously published randomized Phase 1 clinical trial in healthy volunteers [26]. The 1.4 mL per kg mouse dose corresponds to a human equivalent dose (HED) of 0.114 ml/kg of RJX, which is 15% of its MTD determined in the previously published randomized Phase 1 clinical trial in healthy volunteers.

**Patients and Patient Disposition.**

As shown in **Figure S1**, 21 patients were screened. There were 8 screen failures. 6 patients were enrolled in Cohort 1 and 7 patients were enrolled in Cohort 2. One of the Cohort 2 patients withdrew consent on day 4. Consequently, 13 patients were evaluable for safety evaluation and 12 patients were evaluable for efficacy evaluation.

**Patient Evaluations**

An 8-point ordinal scale was used as an assessment of the clinical status at the first assessment of a given study day

***8-point Ordinal Scale:***

1. Death
2. Hospitalized, on invasive mechanical ventilation or ECMO
3. Hospitalized, on non-invasive ventilation or high flow oxygen devices
4. Hospitalized, requiring supplemental oxygen
5. Hospitalized, not requiring supplemental oxygen - requiring ongoing medical care (COVID-19 related or otherwise)
6. Hospitalized, not requiring supplemental oxygen - no longer requires ongoing medical care
7. Not hospitalized, limitation on activities and/or requiring home oxygen
8. Not hospitalized, no limitations on activities

The performance status during follow-up evaluations was examined using the Eastern Cooperative Group (**ECOG**) scale. ECOG score of 0 (i.e., fully active, able to carry on all pre-disease performance without restriction) for outpatients correspond to a health score of 8 on the 8-point ordinal scale. ECOG score of 5 (i.e., dead) corresponds to a score of 1 on the 8-point ordinal sale.

**Supplemental Results**

***Effectiveness of treatment with RJX in a side-by-side comparison to dexamethasone (DEX) in reversing acute lung injury and acute liver injury in mice injected with LPS-GalN***

We first set out to determine the effects of RJX vs. DEX on the LPS-GalN induced inflammatory cytokine response in BALB/c mice (**Fig. S2**). At 2 hours post-injection of LPS-GalN, the serum IL-6 and TNF-α levels were markedly elevated in electively terminated control mice consistent with an inflammatory cytokine response. Low dose RJX at both 0.7 ml/kg (HED: 0.057 ml/kg; 7.5% of human MTD) and 1.4 ml/kg (HED: 0.114 ml/kg; 15% of human MTD) dose levels administered at 2 h after LPS-GalN injection effectively reversed the LPS-GalN induced increased serum levels of the pro-inflammatory cytokines IL-6 and TNF-α within 24 h (Fig. S1). At both of these low dose levels, RJX was significantly more effective than 0.1 mg/kg DEX (HED: 0.008 mg/kg; 0.65 mg for an 80 kg person) or 0.6 mg/kg DEX (HED: 0.05 mg/kg; 4 mg standard dose for an 80 kg person) in reducing the IL-6 levels. 1.4 ml/kg low dose (15% of MTD) RJX was as effective as 6.0 mg/kg supra-therapeutic high dose DEX (HED: 0.49 mg/kg; 39 mg dose for an 80 kg person, which is 4.9-9.8 fold higher than the standard 4-8 mg dose levels for DEX) in reducing the TNF-α levels and slightly less effective in reducing the IL-6 levels (Fig. S2). By comparison, treatment with NS included as vehicle control did not reverse the fulminant cytokine response or prevent its progression.

As evidenced in **Fig. S3** (Panel A) and **Fig. S4**, RJX at 0.7 ml/kg (Mean±SE ALI score: 2.7±0.2) or 1.4 ml/kg (Mean±SE ALI score: 2.3±0.2) low dose levels as well as DEX at the 0.6 mg/kg (HED: 0.05 mg/kg; 4 mg standard dose for an 80 kg person) (Mean±SE ALI score: 2.8±0.3) dose level (but not DEX at 0.1 mg/kg dose level; Mean±SE ALI score: 3.5±0.2) were capable of partially reversing the lung injury that was documented at 2 h post-LPS-GalN injection when treatments were initiated (Mean±SE ALI score: 3.0±0.3), as measured by the lung histopathological scores (i.e., acute lung injury [ALI] scores). The best results were obtained with 6.0 mg/kg supra-therapeutic high dose DEX (HED: 0.49 mg/kg; 39 mg dose for an 80 kg person) (Mean±SE ALI score: 1.8±0.3; Fig. S3A). By comparison, the lung damage further progressed in control mice treated with NS (vehicle) (Mean±SE ALI score: 3.7±01).

Similar to its effects on the LPS-GalN induced ALI, 0.7 ml/kg or 1.4 ml/kg low dose RJX, as well as standard 0.6 mg/kg and very high 6.0 mg/kg dose levels of DEX (but not DEX at 0.1 mg/kg dose level), significantly reduced the liver injury (**Fig. S3; Fig. S5**). The histopathological liver damage scores (Mean ±SE) were 0±0 for control mice not challenged with LPS-GalN, 3.5±0.3 for mice electively terminated 2 h post LPS-GalN, 3.5±0.2 for mice treated with NS post LPS-GalN,

3.3±0.2 for 0.1 mg/kg DEX, 2.7±0.2 for 0.6 mg/kg DEX, 2.3±0.2 for 6.0 mg/kg DEX, 2.5±0.2 for 0.7 ml/kg RJX, and 2.3±0.2 for 1.4 ml/kg RJX.

The observed reversal of the inflammatory cytokine response by RJX was associated with a significant improvement of the survival outcome in this LPS-GalN model of sepsis. Notably, 0.7 ml/kg low dose RJX was moderately more effective than DEX at a 0.1 mg/kg low dose level (Median survival: 15.1 h vs. 5.1 h; 24-h mortality: 50% vs. 83.3%), and it was as effective as DEX at the standard dose 0.6 mg/kg (Fig. S5). Notably, at a dose level of 1.4 ml/kg, which corresponds to 15% of its clinical MTD, RJX reduced the mortality to 40% (Median survival >24h). These results were very similar to and statistically not different from the 33.3% mortality (Median survival >24h) (P=0.99) achieved with DEX at the supratherapeutic 6.0 mg/kg dose level that is 4.9-9.8 fold higher than the standard 4-8 mg clinically applied dose levels for DEX (**Fig. S6**).

*Effectiveness of treatment with RJX in combination with dexamethasone (DEX) in reversing fatal cytokine storm, acute lung injury and acute liver injury in mice injected with LPS-GalN*

Low dose RJX (0.7 ml/kg) plus high dose DEX (6.0 mg/kg) effectively reversed the increased serum levels of the systemic inflammation markers (IL-6, TNF-α, and LDH) within 24 h, and it appeared to be overall more effective than DEX alone or RJX alone (**Fig. S7**). The serum levels of LDH, a biomarker of systemic inflammation and tissue damage as significantly lower in mice treated with the RJX + DEX combination than mice treated with RJX alone (p<0.0001) or DEX alone (p<0.0001) (**Fig. S7**).

As there was significant residual tissue damage in the lungs and liver of surviving LPS-GalN challenged mice treated with RJX or DEX (even at the 6.0 mg/kg high dose level), we next sought to determine if a combination of low dose RJX (0.7 ml/kg) and high dose DEX (6.0 mg/kg) could improve the tissue healing and the survival outcome after LPS-GalN exposure. Treatments were initiated at 2 h after LPS-GalN injection at a time of documented active systemic inflammation.

LPS-GalN caused severe inflammation and oxidative stress in the lungs and liver with enhanced lipid peroxidation, as measured by a marked elevation of MDA levels, and decreased tissue levels of Vitamin C as well as the anti-oxidant enzymes GSH-Px that inhibits lipid peroxidation and SOD that converts superoxide anion radicals that contribute to lipid peroxidation into hydrogen peroxide and oxygen. Low dose RJX alone or in combination with DEX significantly suppressed the oxidative stress, as documented by increases in Vitamin C, GSH-Px and SOD levels that were reduced by LPS-GalN (**Figure S8**). Notably, delayed treatments with RJX, DEX or RJX+DEX starting at 2 h after the LPS-GalN injection partially reversed the lung damage, as evidenced by significantly reduced histopathological lung scores (**Fig. S9, Fig. S10**). The tissue healing activity of the combination was more pronounced than the tissue healing activity of RJX alone or DEX alone (Fig. S4, Fig. S5). The lung histopathological ALI scores ranged from 3 to 4 for the LPS-GalN + NS, from 2-3 for the LPS-GalN+RJX, from 1-3 for LPS-GalN + DEX, and from 1-2 for LPS-GalN + RJX + DEX. Hence, although treatments were delayed until the onset of fulminant cytokine storm and systemic inflammation with severe oxidative stress as well as very severe lung damage, a near-complete recovery of the inflammatory lung injury was achieved within 24 h.

Similar to its effects on the LPS-GalN induced ALI, RJX + DEX combination significantly reduced the liver injury (**Fig. S9**). While 12 of 12 mice (100%) treated with either RJX alone (N=6) or DEX alone (N=6) had moderate to severe residual damage in either their lungs or liver, 2 of the 6 mice (33.3%) treated with the combination regimen had no or minimal damage (i.e., histopathological damage scores: 0-1) in both organs ($p$=0.098, Fisher's exact test).

The combination therapy was more effective than RJX alone or DEX alone in improving the survival outcome (**Fig. S11**). In contrast to the rapid death of all control mice treated with NS (Median survival: 4.3 h post-LPS-GalN or 2.3 h after administration of NS), 100% of mice treated with RJX + DEX survived the LPS-GalN challenge (Median survival: >24 hours post LPS-GalN or >22 hours after initial administration of RJX+DEX) (Fig. S2). By comparison, the combined group of mice treated with monotherapy (i.e., RJX alone or DEX alone) (n=12) had a 24-hour survival rate of 41.7% (Monotherapy with RJX or DEX vs. Combination therapy with RJX+DEX: Log-rank $X^2$ = 3.053, $p$=0.081).

#### SUPPLEMENTAL FIGURES

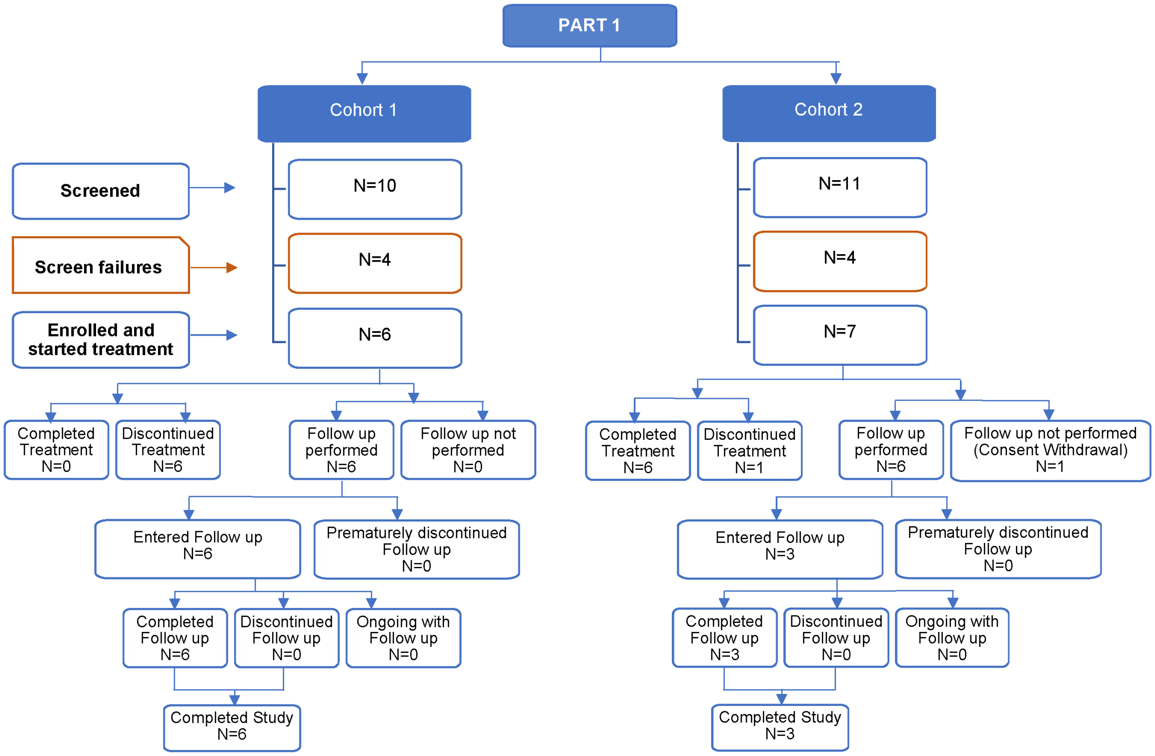


**Figure S1. Patient Disposition**

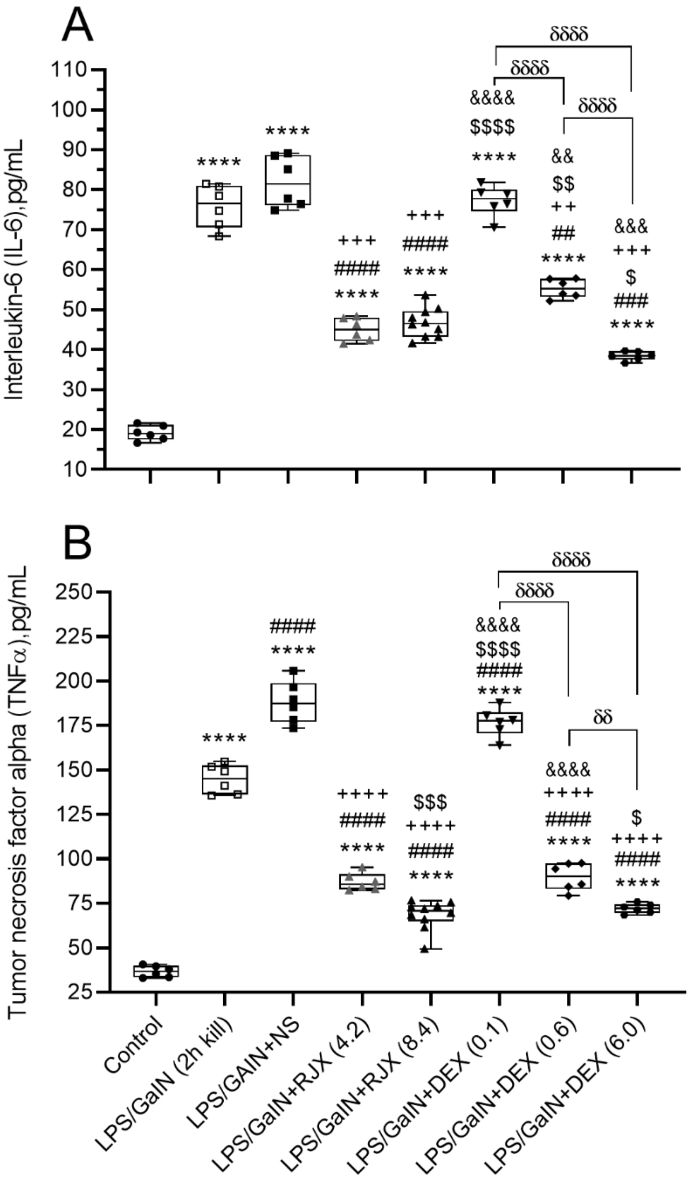


**Fig. S2.** The effects of Rejuveinix (RJX) and the different doses of the Dexamethasone (DEX), treatments on serum interleukin 6 (IL-6; Panel A), tumor necrosis factor-alpha (TNF-α; Panel B) in a Mouse Model of Fatal Cytokine Storm, Sepsis, Systemic Inflammation, ARDS and Multiorgan Failure. BALB/C mice were treated with i.p injections of RJX (0.7 ml/kg = 4.2 ml/kg of 6-fold diluted RJX, 0.5 ml/mouse; or 1.4 ml/kg = 8.4 ml/kg of 1:6-diluted RJX, 0.5 ml/mouse), DEX (0.1 mg/kg, 0.6 mg/kg and 6.0 mg/kg), or vehicle (NS, 0.5 mL/mouse) two hours post-injection of LPS-GalN, or terminated at 2 hours post LPS-GalN injection without any therapeutic intervention. Except for untreated control mice (Control), each mouse received 0.5 ml of LPS-GalN (consisting of 100 ng of LPS plus 8 mg of D-galactosamine i.p.). The depicted Whisker plots represent the median and values for serum IL-6 and TNF-α levels from all 6 mice from each group except for the 1.4 ml/kg RJX group where blood samples were obtained from all 10 mice. In Panel A, Welch's ANOVA and Tamhane's T2 post-hoc test were used for comparing the results among different treatment groups. In Panel B, ANOVA and Tukey post-hoc test were used for comparing the results among different treatment groups. Statistical significance between groups is shown by **** $p<0.0001$ as compared to control group, and ## $p<0.01$; ### $p<0.001$; #### $p<0.0001$ as compared to LPS/GalN (2h kill) group, ++ $p<0.01$; +++ $p<0.001$; ++++ $p<0.0001$ as compared to LPS/GalN+NS group, \$ $p<0.05$; \$\$ $p<0.01$; \$\$\$ $p<0.001$; \$\$\$\$ $p<0.0001$ LPS/GalN+RJX (4.2) group, && $p<0.01$; &&& $p<0.001$; &&&& $p<0.0001$ as compared to LPS/GalN+RJX (8.4) group, and and δδ $p<0.01$; δδδδ $p<0.0001$ pairwise comparison.

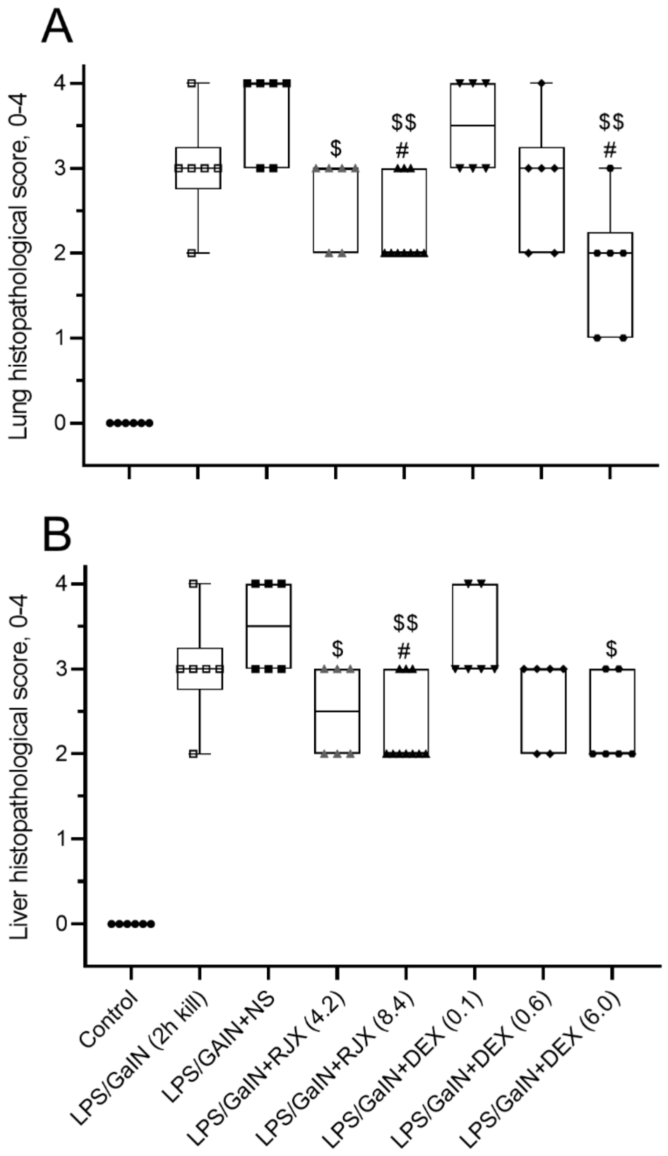


**Fig. S3.** Tissue-Level In Vivo Activity of Rejuveinix (RJX) and the different doses of the Dexamethasone (DEX), treatments on Lung and Liver Histopathological Scores in a Mouse Model of Fatal Cytokine Storm, Sepsis, Systemic Inflammation, ARDS and Multiorgan Failure. BALB/C mice were treated with i.p injections of RJX (6-fold diluted, 4.2 mL/kg or 8.4 mL/kg, 0.5 ml/mouse), DEX (0.1 mg/kg, 0.6 mg/kg and 6.0 mg/kg), or vehicle (NS, 0.5 mL/mouse) two hours post-injection of LPS-GalN, and only LPS-GalN for 2 hours. Except for untreated control mice (Control), each mouse received 0.5 ml of LPS-GalN (consisting of 100 ng of LPS plus 8 mg of D-galactosamine i.p.). The depicted Whisker plots represent the median and values. In (A), the lung histopathological score ("lung injury score") was graded according to a 5-point scale from 0 to 4 as follows: 0, l, 2, 3, and 4 represented no damage, mild damage, moderate damage, severe damage, and very severe damage, respectively. In (B), the liver histopathological score (“liver injury score”) was graded according to a 5-point scale from 0 to 4 as follows: 0, l, 2, 3, and 4 represented no damage, mild damage, moderate damage, severe damage, and very severe damage, respectively. Kruskal-Wallis test and Mann Whitney U test were used for comparing the results among different treatment groups. Statistical significance between groups is shown by # $p<0.05$; as compared to LPS/GaIN (2h kill) group, and $ $p<0.05$; $$ $p<0.01$; as compared to LPS/GaIN+NS group.

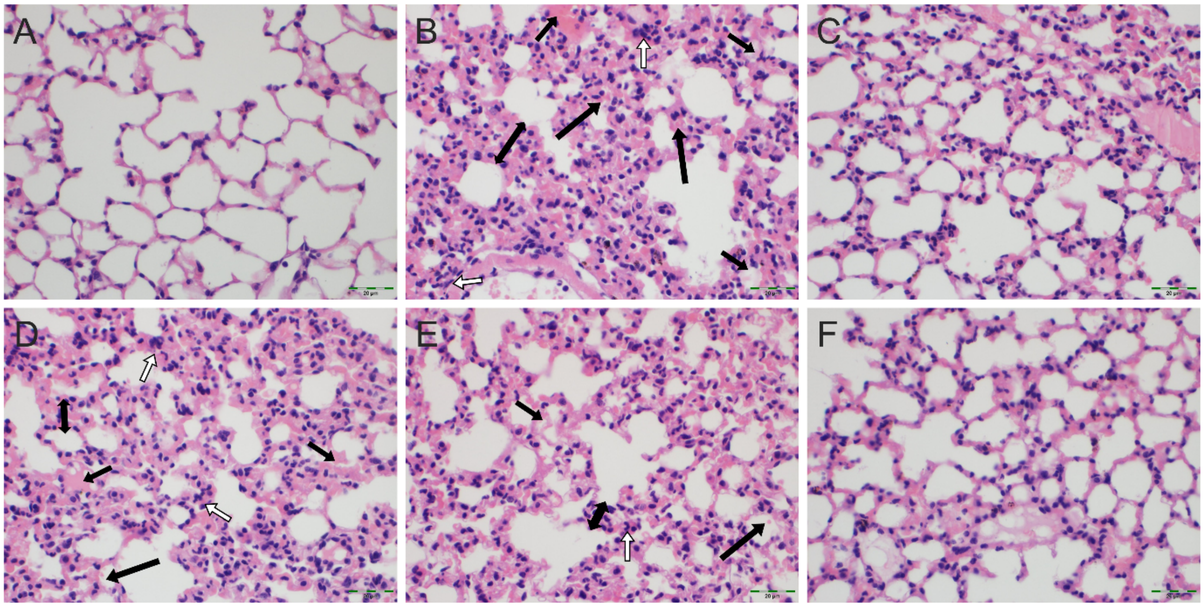


**Fig. S4.** The Effects of Rejuveinix (RJX) and the different doses of the Dexamethasone (DEX), treatments on Acute Lung Injury and Inflammation in a Mouse Model of Fatal Cytokine Storm, Sepsis, Systemic Inflammation, ARDS and Multiorgan Failure. Groups of 6 BALB/C mice were treated with i.p injections of RJX (6-fold diluted, 4.2 mL/kg, 0.5 ml/mouse), DEX (0.1 mg/kg, 0.6 mg/kg, and 6.0 mg/kg, 0.5 mL/mouse), or vehicle (NS, 0.5 mL/mouse) two hours post-injection of LPS-GalN. Except for untreated control mice (Control, Panel A), each mouse received 0.5 ml of LPS-GalN (consisting of 100 ng of LPS plus 8 mg of D-galactosamine i.p.). The lung histopathological ALI scores were 0 for each of the control mice (Panel A), ranged from 3 to 4 (Median: 4) for the LPS-GalN+NS group (Panel B), from 2-3 (Median: 3) for LPS-GalN+RJX (0.7 ml/kg) group (Panel C), from 3-4 (Median: 3.5) for LPS-GalN+DEX (0.1 mg/kg) (Panel D), from 2-4 (Median: 3) for LPS-GalN+DEX (0.6 mg/kg) group (Panel E) and from 1-3 (Median: 2) for LPS-GalN+DEX (6.0 mg/kg) group (Panel F). Depicted are microscopic images of lung tissues of representative mice from the untreated control group and various treatment groups. White arrow: inflammatory cell infiltration; Black arrow (short): Exudate, edema; Black arrow (Long): hemorrhage; Black double-headed arrow: the thickness of the alveolar wall. H&E X400.

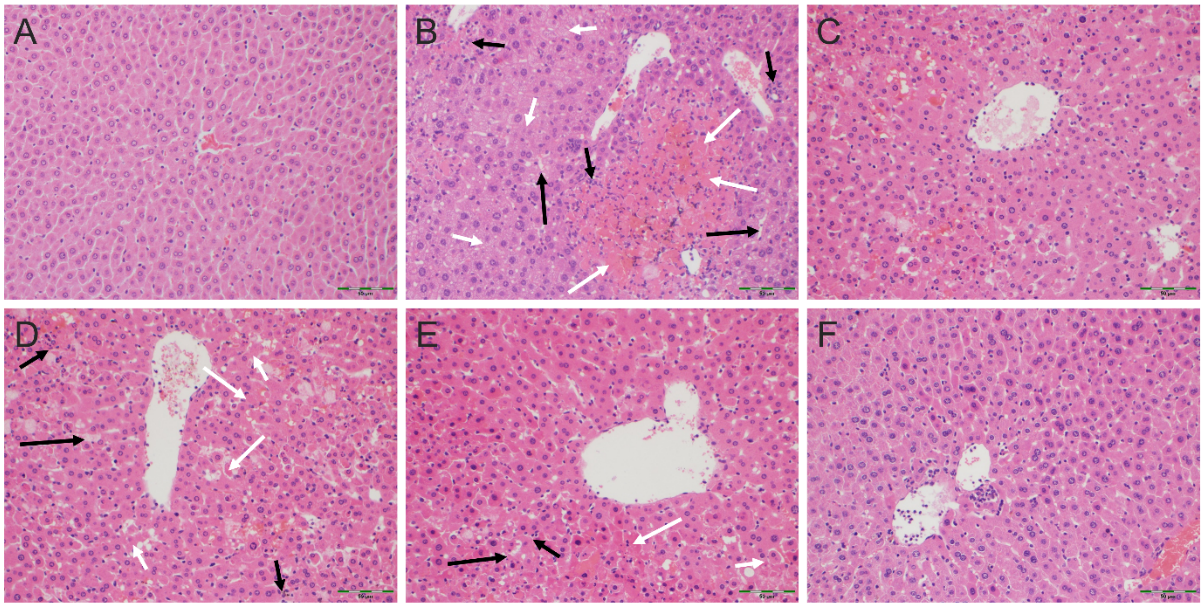


**Fig. S5.** The Effects of Rejuveinix (RJX) and the different doses of the Dexamethasone (DEX), treatments on Liver Injury and Inflammation in a Mouse Model of Fatal Cytokine Storm, Sepsis, Systemic Inflammation, ARDS and Multiorgan Failure. Groups of 6 BALB/C mice were treated with i.p injections of RJX (6-fold diluted, 4.2 mL/kg, 0.5 ml/mouse), DEX (0.1 mg/kg, 0.6 mg/kg, and 6.0 mg/kg, 0.5 mL/mouse), or vehicle (NS, 0.5 mL/mouse) two hours post-injection of LPS-GalN. Except for untreated control mice (Control Panel A), each mouse received 0.5 ml of LPS-GalN (consisting of 100 ng of LPS plus 8 mg of D-galactosamine i.p.). The liver histopathological scores ranged from 3 to 4 for the LPS-GalN+NS group (Panel B), from 2-3 for LPS-GalN+RJX group (Panel C), from 3-4 for LPS-GalN+DEX (0.1) (Panel D), from 2-3 for LPS-GalN+DEX (0.6) group (Panel E) and from 2-3 for LPS-GalN+DEX (6.0) group (Panel F). Depicted are microscopic images of the liver tissues of representative mice from the untreated control group and various treatment groups. While the liver histopathological score median was 0 for untreated control mouse, the liver score median for the depicted mice treated with 0.6 mg/kg or 6.0 mg/kg DEX were 3 for each, and the liver score median for the LPS-GaIN+NS treated control mice was 3.5. Black arrow (short): Inflammatory cell infiltration; Black arrow (long): Congestion; White arrow (long): Necrosis; White arrow (short): Hydropic degeneration. H&E 200x

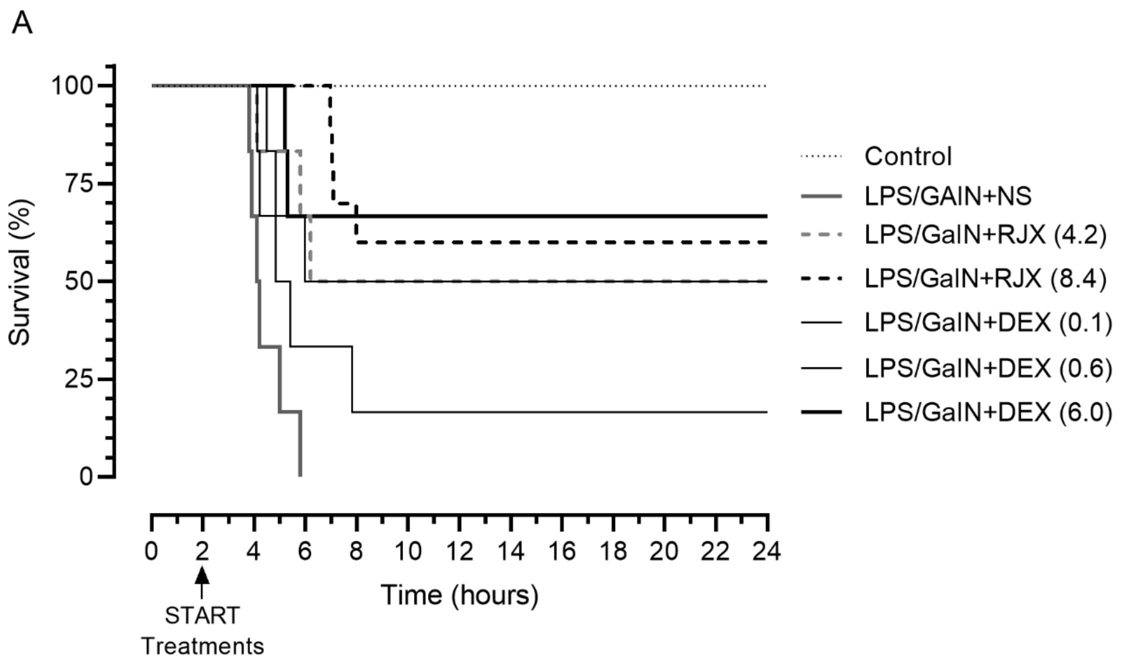


| **B. Survival Analysis** | Censored | Events | n | Proportion | Median |
|---|---|---|---|---|---|
| Control | 6 | 0 | 6 | 0.0 % | ≥24 h |
| LPS/GaIN+NS | 0 | 6 | 6 | 100.0 % | 4.2 h |
| LPS/GaIN +RJX (4.2) | 3 | 3 | 6 | 50.0 % | 15.1 h |
| LPS/GaIN +RJX (8.4) | 6 | 4 | 10 | 40.0 % | ≥24 h |
| LPS/GaIN +DEX (0.1) | 1 | 5 | 6 | 83.3 % | 5.1 h |
| LPS/GaIN +DEX (0.6) | 3 | 3 | 6 | 50.0 % | 15.0 h |
| LPS/GaIN +DEX (6.0) | 4 | 2 | 6 | 33.3 % | ≥24 h |

| **C. Pairwise comparisons** | | Log-rank Chi-square | *p* value | *p* value summary |
|---|---|---|---|---|
| LPS/GaIN+NS | LPS/GaIN+RJX(4.2) | 6.669 | 0.0098 | ** |
| LPS/GaIN+NS | LPS/GaIN+RJX(8.4) | 19.259 | 0.0001 | **** |
| LPS/GaIN+NS | LPS/GAIN+DEX (0.1) | 2.401 | 0.1213 | ns |
| LPS/GaIN+NS | LPS/GAIN+DEX (0.6) | 6.066 | 0.0138 | * |
| LPS/GaIN+NS | LPS/GAIN+DEX (6.0) | 8.044 | 0.0046 | ** |
| LPS/GaIN+RJX(4.2) | LPS/GAIN+DEX (0.1) | 1.378 | 0.2405 | ns |
| LPS/GaIN+RJX(4.2) | LPS/GAIN+DEX (0.6) | 0.004 | 0.9467 | ns |
| LPS/GaIN+RJX(4.2) | LPS/GAIN+DEX (6.0) | 0.193 | 0.6606 | ns |
| LPS/GaIN+RJX(8.4) | LPS/GaIN+DEX (0.1) | 5.288 | 0.0215 | * |
| LPS/GaIN+RJX(8.4) | LPS/GaIN+DEX (0.6) | 0.633 | 0.4264 | ns |
| LPS/GaIN+RJX(8.4) | LPS/GaIN+DEX (6.0) | 0.001 | 0.9991 | ns |
| LPS/GaIN+DEX (0.1) | LPS/GaIN+DEX (0.6) | 1.220 | 0.2694 | ns |
| LPS/GaIN+DEX (0.1) | LPS/GaIN+DEX (6.0) | 2.848 | 0.0915 | ns |

**Fig. S6.** *In Vivo* Treatment Activity of Rejuveinix (RJX) and the different doses of the Dexamethasone (DEX) in the LPS-GalN Mouse Model of Fatal Cytokine Storm, Sepsis, Systemic Inflammation, ARDS and Multiorgan Failure. BALB/C mice were treated with i.p injections of RJX (4.2 mL/kg or 8.4 mL/kg of 1:6-diluted RX = 0.7 mL/kg or 1.4 mL/kg RJX; 0.5 ml/mouse), DEX (0.1 mg/kg, 0.6 mg/kg and 6.0 mg/kg 0.5 mL/mouse), or vehicle (NS, 0.5 mL/mouse) two hours post-injection of LPS-GalN. Except for untreated control mice (Control), each mouse received 0.5 ml of LPS-GalN (consisting of 100 ng of LPS plus 8 mg of D-galactosamine i.p.). The cumulative proportion of mice remaining alive (Survival, %) is shown as a function of time after the LPS-GalN challenge. Depicted are the Kaplan Meier survival curves (Panel A) and survival data with statistical analysis (Panels B and C) of the different treatment groups.

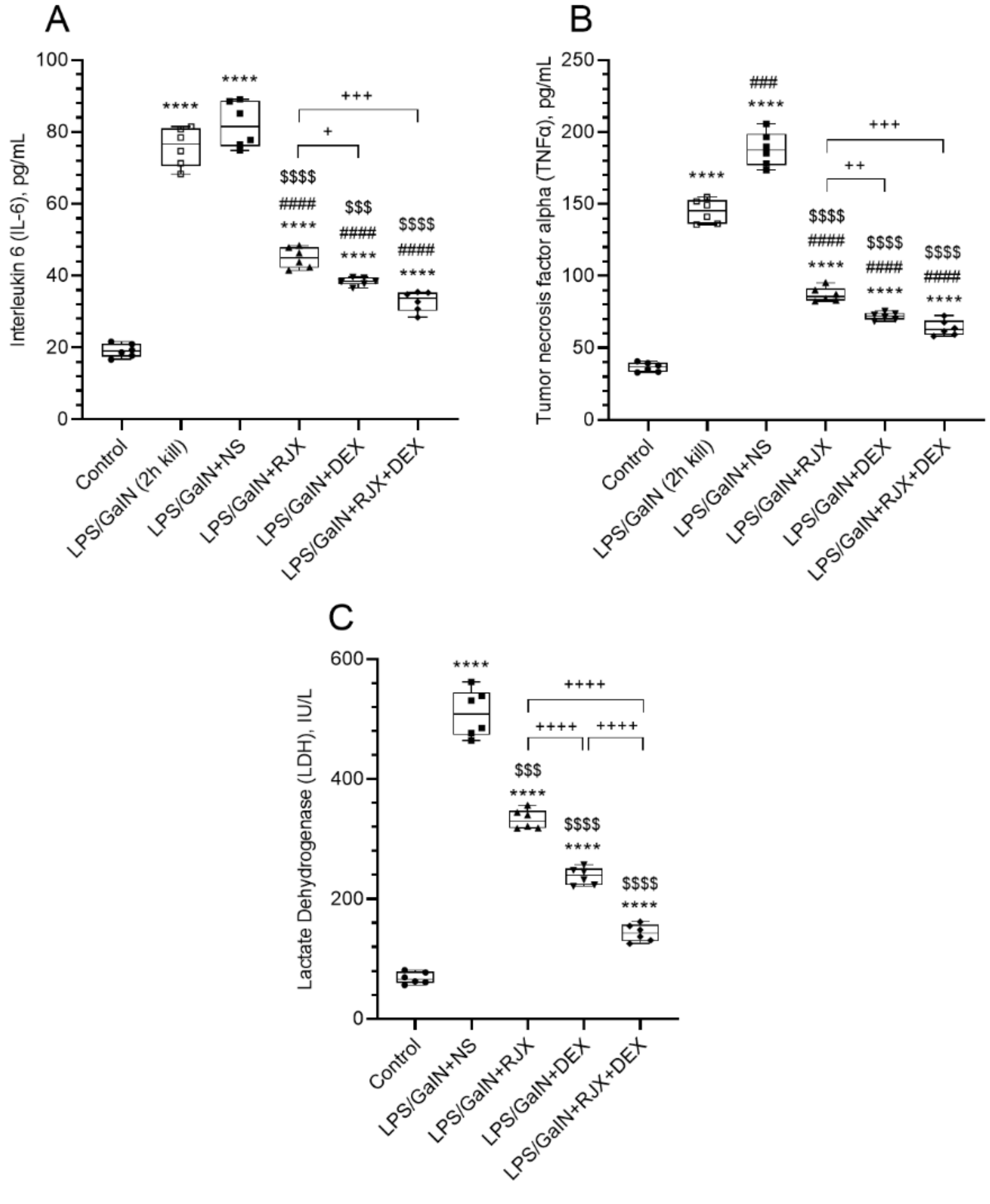


**Fig. S7.** Therapeutic Use of Low Dose RJX Plus Supratherapeutic High Dose DEX Combination after Onset of Systemic Inflammation and Lung Injury Reverses Inflammatory Cytokine Response and Systemic Inflammation in the LPS-GalN Mouse Model of Fatal Cytokine Storm and Sepsis. Depicted are the effects of Rejuveinix (RJX), Dexamethasone (DEX), and RJX + DEX combination treatments on serum levels of interleukin 6 (IL-6; Panel A), tumor necrosis factor-alpha (TNF-α; Panel B), and lactate dehydrogenase (LDH; Panel C). Groups of 6 BALB/C mice were treated with i.p injections of RJX (6-fold diluted, 4.2 mL/kg, 0.5 ml/mouse), DEX (6 mg/kg, 0.5 mL/mouse), RJX + DEX (0.5 ml/mouse), or vehicle (NS, 0.5 ml/mouse) two hours post-injection of LPS-GalN. Except for untreated control mice (Control), each mouse received 0.5 ml of LPS-GalN (consisting of 100 ng of LPS plus 8 mg of D-galactosamine) i.p. The depicted Whisker plots represent the median and values. Welch's ANOVA and Tamhane's T2 post-hoc test was used for comparing the results among different treatment groups. Statistical significance between groups is shown by **** p<0.0001 as compared to control group, ### p<0.001; #### p<0.0001 as compared to LPS/GaIN (2h kill) group, $$$ $p<0.001$; $$$$ $p<0.0001$ as compared to LPS/GaIN+NS, and + $p<0.05$; ++ $p<0.01$; +++ $p<0.001$; ++++ $p<0.0001$ pairwise comparisons between the groups.

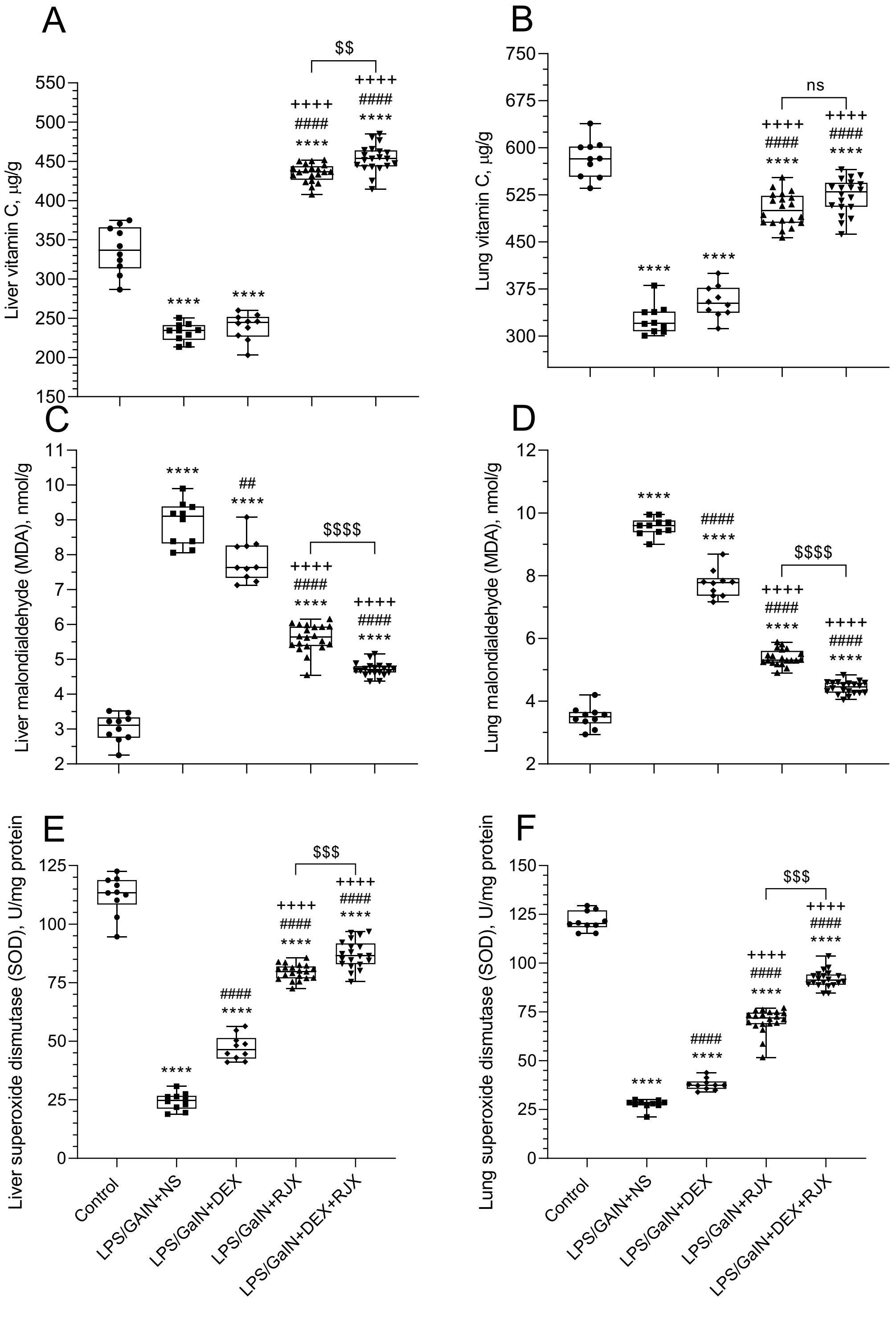


**Figure S8. Tissue-Level In Vivo Anti-Oxidant Activity of Rejuveinix (RJX), Dexamethasone (DEX), and RJX+DEX in a Mouse Model of Fatal Cytokine Storm, Sepsis, Systemic Inflammation, ARDS and Multiorgan Failure.** BALB/C mice were treated with i.p injections of RJX (n=20, 6-fold diluted, 4.2 mL/kg, 0.5 ml/mouse), DEX (n= 10, 6 mg/kg, 0.5 mL/mouse), RJX + DEX (n=20, 0.5 mL/mouse), or vehicle (NS,

0.5 mL/mouse) two hours post-injection of LPS-GalN. Except for untreated control mice (Control, n=10), each mouse received 0.5 ml of LPS-GalN (consisting of 100 ng of LPS plus 8 mg of D-galactosamine) i.p. The depicted Whisker plots represent the median and values. In A, C, E and F, Welch's ANOVA and Games-Howell post-hoc test were used for comparing the results among different treatment groups. In B and D, ANOVA and Tukey's post-hoc test were used for comparing the results among different treatment groups. Statistical significance between groups is shown by **** *p*<0.0001 as compared to Control group, ## *p*<0.01; #### *p*<0.0001 as compared to LPS/GalN+NS group, ++++ *p*<0.0001 as compared to LPS/GalN+DEX group, and $$ *p*<0.01; $$$ *p*<0.001; $$$$ *p*<0.0001 pairwise comparisons between the groups.

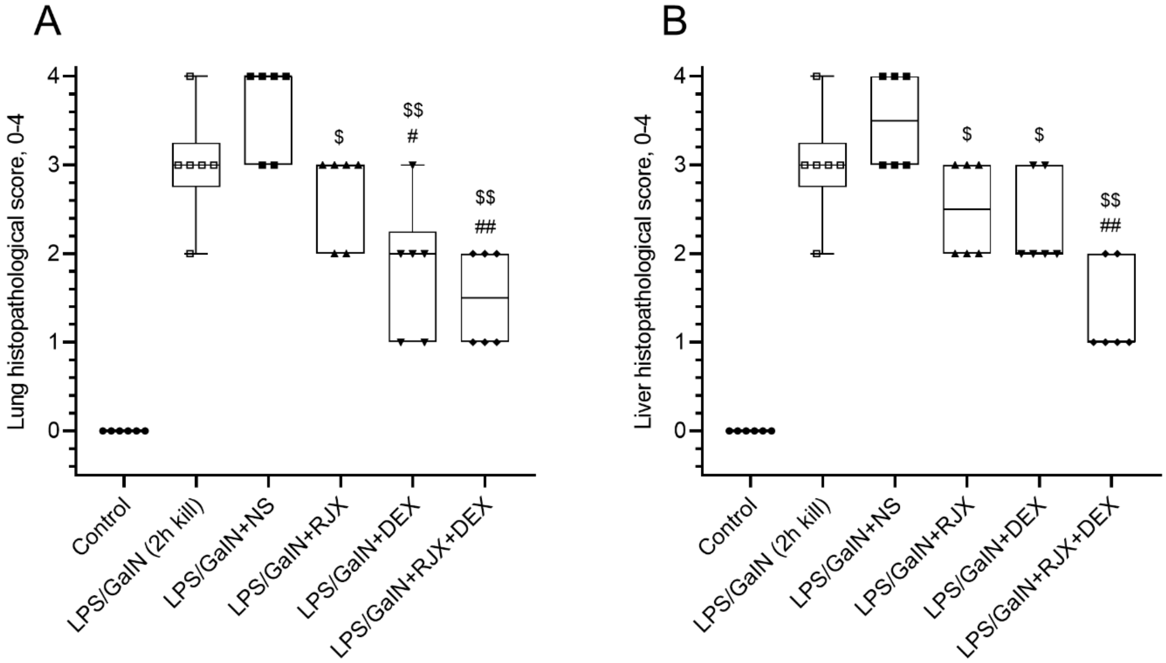


**Fig. S9.** *In Vivo* Treatment Activity of Low Dose RJX, Supratherapeutic High Dose DEX and Their Combination on Lung and Liver Histopathological Scores in the LPS-GalN Mouse Model of Fatal Cytokine Storm and Sepsis. Groups of 6 BALB/C mice were treated with i.p injections of RJX (6-fold diluted, 4.2 mL/kg, 0.5 ml/mouse), DEX (6.0 mg/kg, 0.5 mL/mouse), or vehicle (NS, 0.5 mL/mouse) two hours post-injection of LPS-GalN. Except for untreated control mice (Control), each mouse received 0.5 ml of LPS-GalN (consisting of 100 ng of LPS plus 8 mg of D-galactosamine) i.p. In (A), the lung histopathological score ("lung injury score") was graded according to a 5-point scale from 0 to 4 as follows: 0, l, 2, 3, and 4 represented no damage, mild damage, moderate damage, severe damage, and very severe damage, respectively. In (B), the liver histopathological score ("liver injury score") was graded according to a 5-point scale from 0 to 4 as follows: 0, l, 2, 3, and 4 represented no damage, mild damage, moderate damage, severe damage, and very severe damage, respectively. Mann Whitney U test were used for comparing the results among different treatment groups Statistical significance between groups is shown by # p<0.05; ## p<0.01 as compared to LPS/GaIN (2h kill) group, $ p<0.05; $$ p<0.01 as compared to LPS/GaIN+NS group.

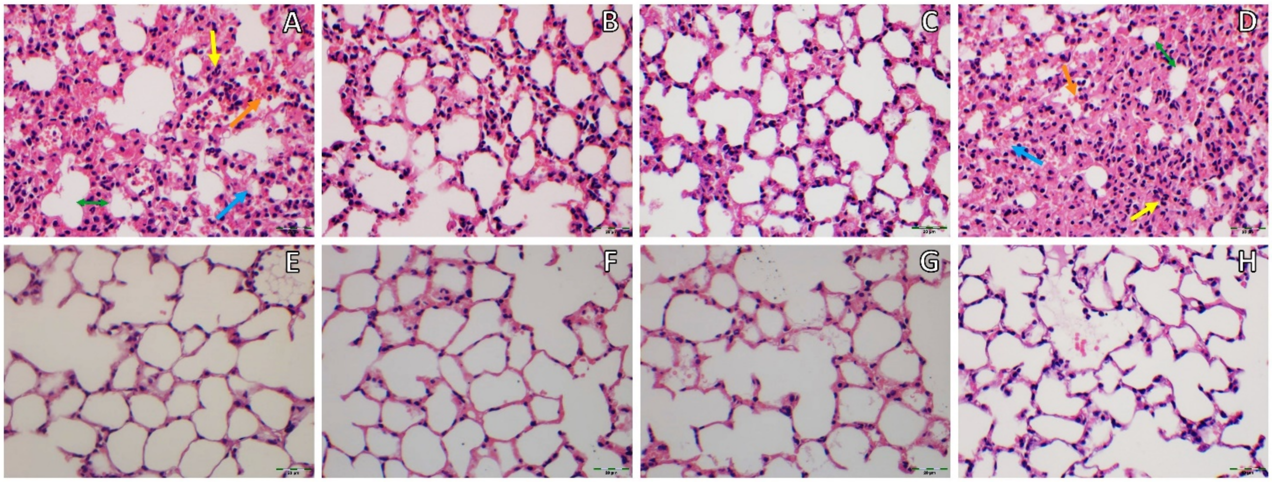


**Fig. S10.** RJX (0.7 mL/kg) plus DEX (6.0 mg/kg) Combination Mitigates Acute Lung Injury and Inflammation in a Mouse Model of Fatal Cytokine Storm and Sepsis. [A]: Lung tissue of a representative mouse injected with LPS-GalN without any pre- or post-LPS-GalN treatments and electively sacrificed at 2 hours to confirm the rapid onset of lung damage. The histopathological ALI score was 3 consistent with severe lung damage. Yellow arrow: inflammatory cell infiltration; blue arrow: exudate; orange arrow: hemorrhage; green block: thickness of alveolar wall. [B] Lung tissue of a LPS-GalN injected representative mouse treated with a single dose of RJX at 2 hours post-LPS-GalN. The mouse was electively sacrificed at 24 hours post LPS-GalN. ALI score = 2 (moderate lung damage). [C] Lung tissue of a LPS-GalN injected representative mouse treated with a single dose of DEX at 2 hours post-LPS-GalN. The mouse was electively sacrificed at 24 hours post LPS-GalN. ALI score = 2 (moderate lung damage). [D] ] Lung tissue of a LPS-GalN injected representative mouse treated with a single dose of NS at 2 hours post-LPS-GalN. The mouse died of sepsis at 4.2 hours post LPS-GalN. ALI score = 4 (very severe lung damage). Yellow arrow: inflammatory cell infiltration; blue arrow: exudate; orange arrow: hemorrhage; green block: thickness of alveolar wall. [E] Lung tissue of a healthy control mouse not injected with LPS-GalN and electively sacrificed at 24 hours. ALI score = 0 (no lung damage). [F] & [G]. Lung tissues from two representative LPS-GalN injected control mice treated RJX + DEX 2 hours before at 2 hours post-LPS-GalN. These mice survived the LPS-GalN challenge and were electively sacrificed at 24 hours. No lung damage was detected (Histopathological lung score/ALI score = 0). [H] Lung tissue of a LPS-GalN injected representative mouse treated with RJX + DEX at 2 hours post-LPS-GalN. The mouse was electively sacrificed at 24 hours post LPS-GalN. ALI score = 1 (mild lung damage). H&E X400

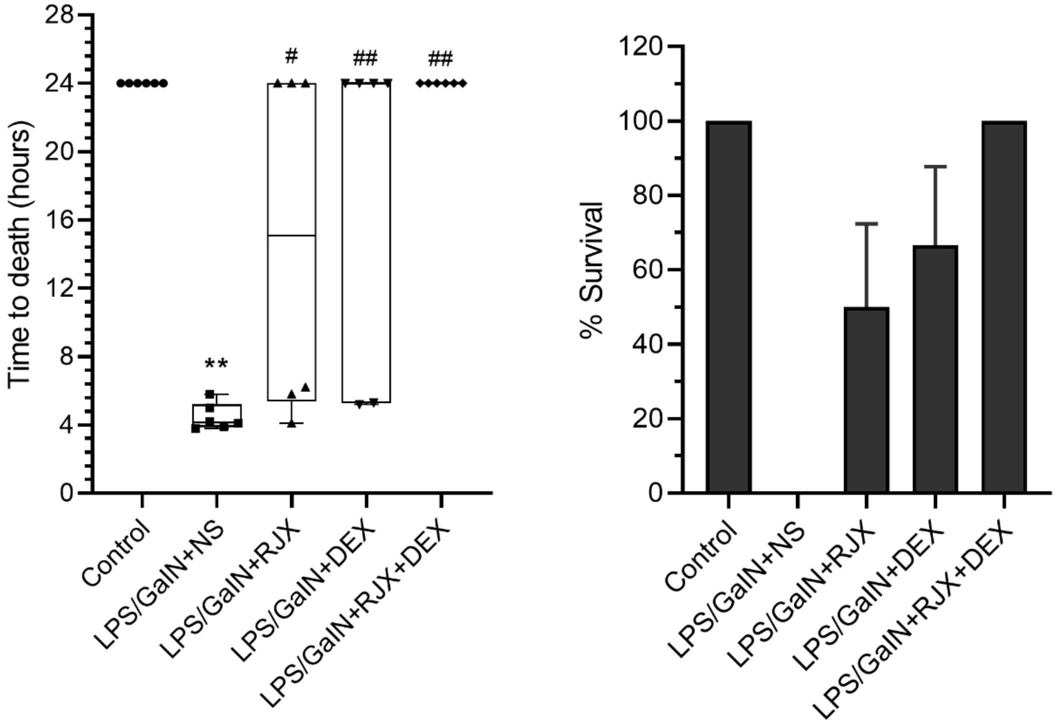


**Fig. S11.** Therapeutic Use of Low Dose RJX + Supratherapeutic High Dose DEX Combination After Onset of Systemic Inflammation and Lung Injury Improves the Survival Outcome in the LPS-GalN Mouse Model of Fatal Cytokine Storm and Sepsis. Groups of 6 BALB/C mice were treated with i.p injections of RJX (6-fold diluted, 4.2 mL/kg, 0.5 ml/mouse), DEX (6 mg/kg, 0.5 mL/mouse), RJX + DEX (0.5 mL/mouse), or vehicle (NS, 0.5 mL/mouse) two hours post-injection of LPS-GalN. Except for untreated control mice (Control), each mouse received 0.5 ml of LPS-GalN (consisting of 100 ng of LPS plus 8 mg of D-galactosamine) i.p. The depicted Whisker plots represent the median and values for time to death from 6 mice in each group. Kruskall Wallis and Mann Whitney U (pairwise comparisons test) test were used for comparing the results among different treatment groups. Statistical significance between groups is shown by: ** $p<0.01$ compared as control group and, # $p<0.05$; ## $p<0.01$ compared as LPS/GaIN+NS group.

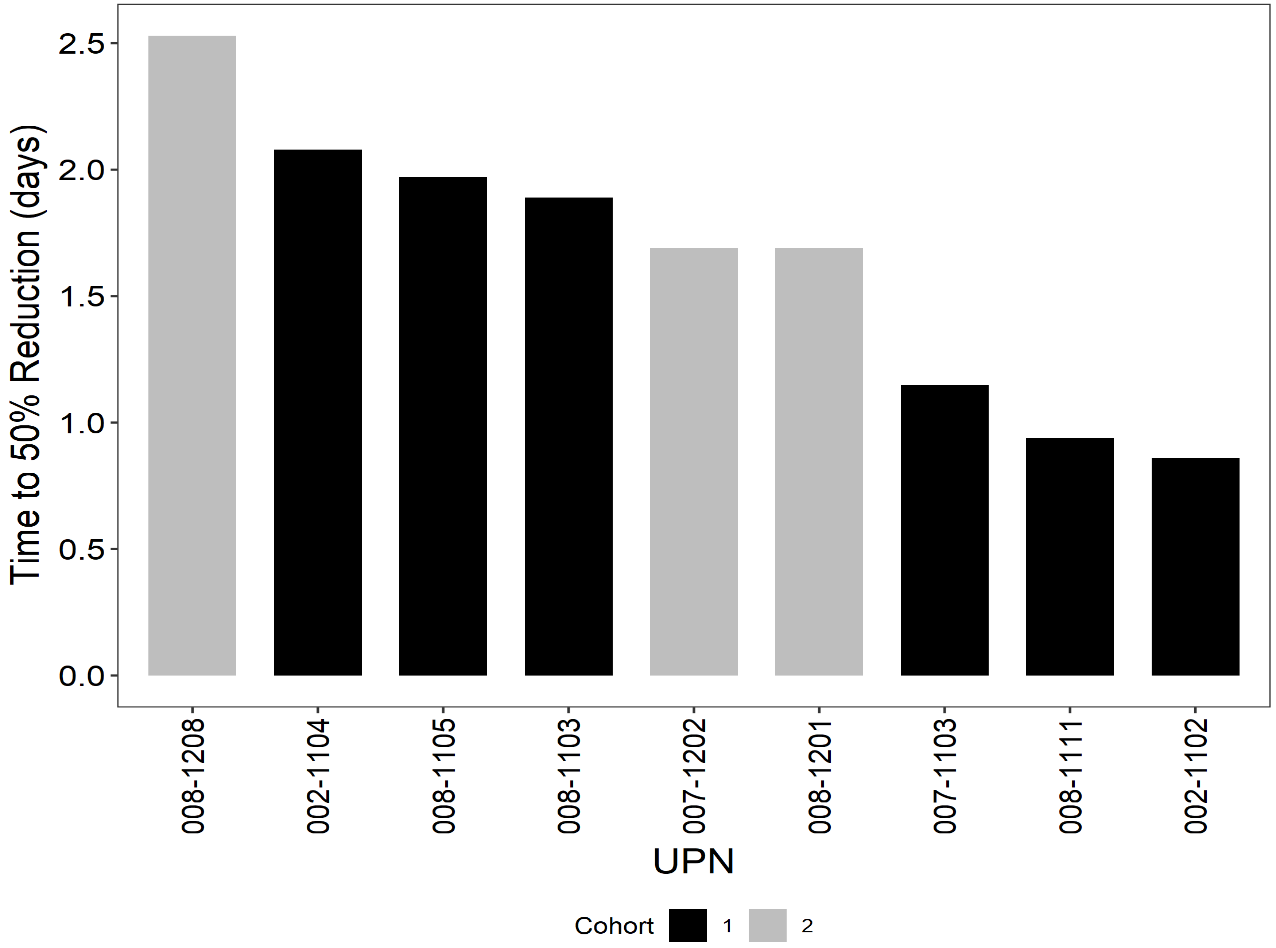


**Figure S12. Swimmer Plot of CRP Normalization Kinetics in Severe COVID-19 Patients Treated with RJX Plus SOC.** Among the 9 patients who recovered after protocol therapy, the estimated time to reduction of baseline CRP values by 50% ranged from 0.9 days to 2.5 days (Median: 1.7 days; Mean±SE = 1.6 ± 0.2 days).

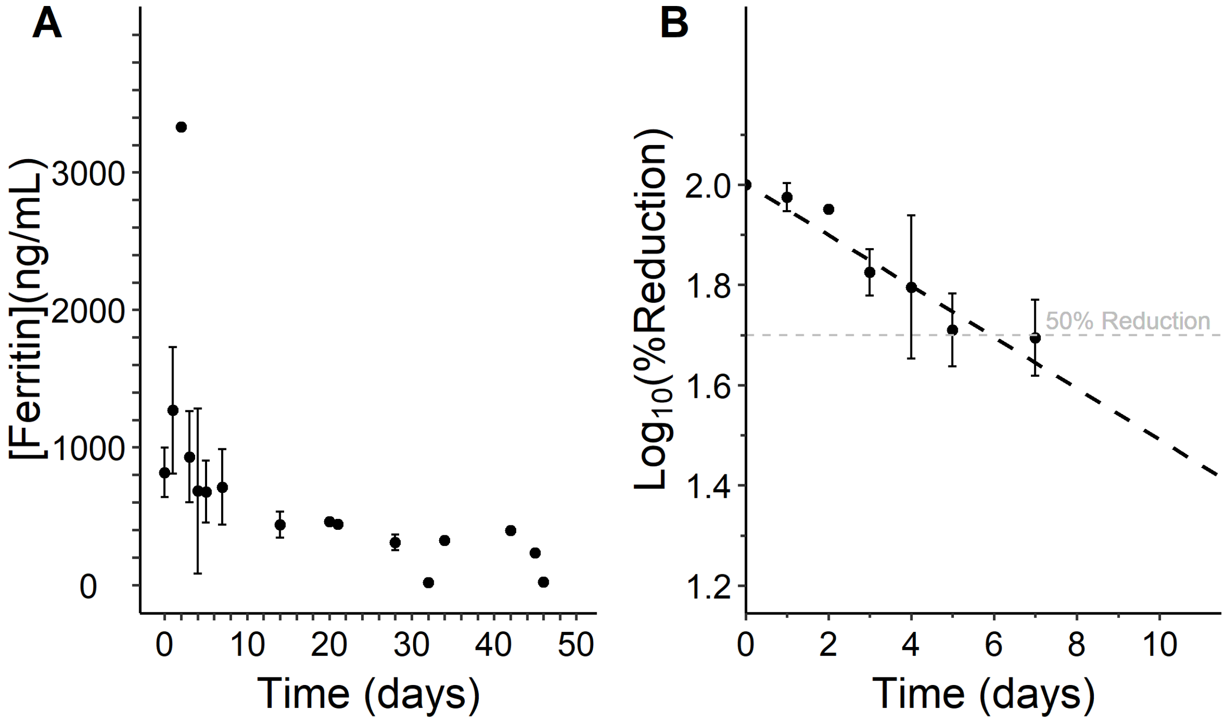


**Figure S13. Time-dependent decrease of Serum Ferritin Values (in ng/mL) in RJX-Treated Severe COVID-19 Patients.** [A] Serum Ferritin concentration is depicted over 46 days for 8 evaluable responders [B] The first order kinetics of the reductions in serum Ferritin values was investigated by fitting a straight line to a semi-log plot of the portion of the Ferritin concentration x time curve that displayed maximum reduction in Ferritin values over the course of RJX treatment. There were 32 independent data points across 7 days. The slope of the line represents the rate constant for Ferritin reduction in $\log_{10}$ scale (viz.: -0.0511). The time to 50% reduction ($T_{50}$ = 5.9 days) of serum Ferritin values was calculated using the rate constant and the intercept value of 2.0. Equation of the line: $\text{Log}_{10}$ (% Reduction) = -0.05107 x Time(Days) + 2.0.

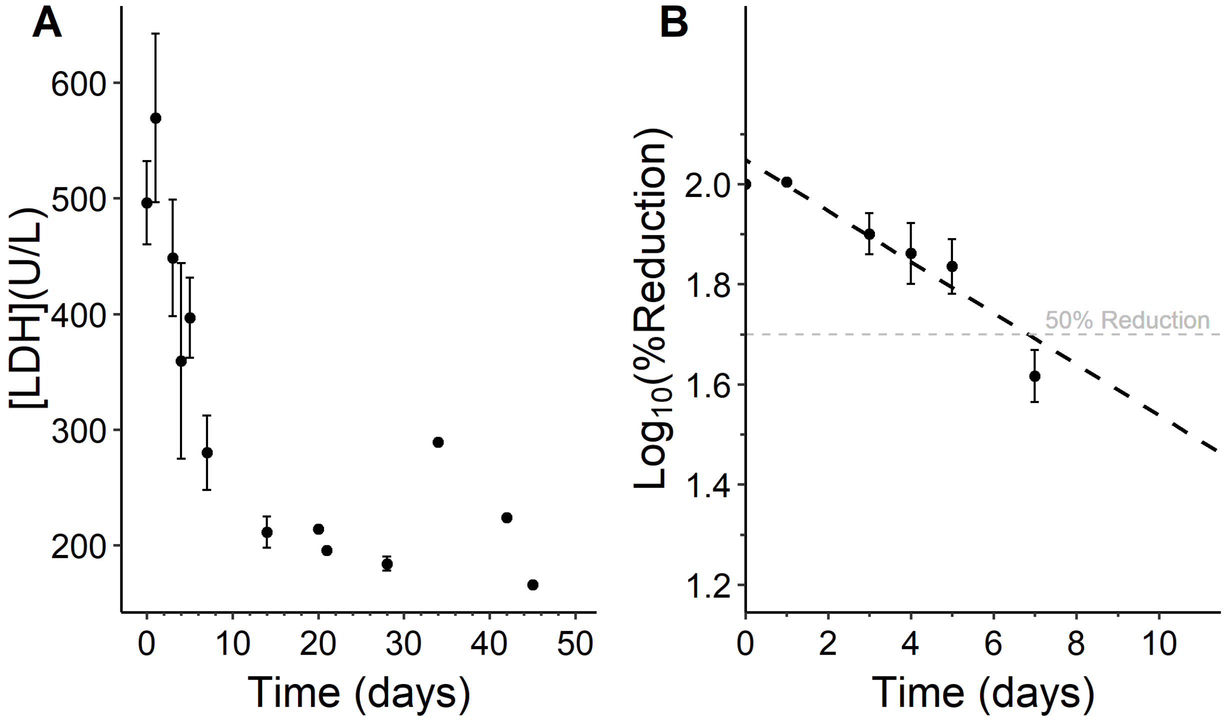


**Figure S14. Time-dependent decrease of Serum LDH Values (in U/L) in RJX-Treated Severe COVID-19 Patients.** [A] Serum LDH concentration is depicted over 46 days for 5 evaluable responders [B] The first order kinetics of the reductions in serum LDH values was investigated by fitting a straight line to a semi-log plot of the portion of the LDH concentration x time curve that displayed maximum reduction in LDH values over the course of RJX treatment. There were 20 independent data points across 7 days. The slope of the line represents the rate constant for LDH reduction in $\log_{10}$ scale (viz.: -0.0511). The time to 50% reduction ($T_{50}$ = 6.9 days) of serum LDH values was calculated using the rate constant and the intercept value of 2.05. Equation of the line: $\text{Log}_{10}$ (% Reduction) = -0.0511 x Time(Days) + 2.05.

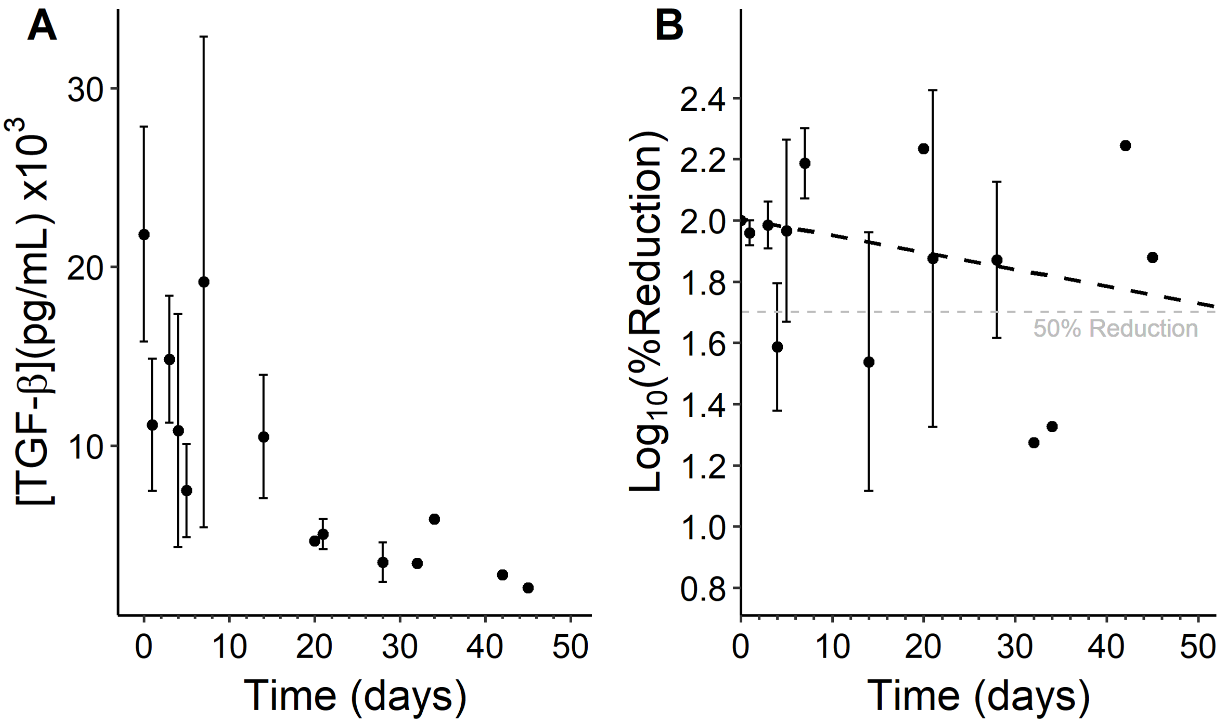


**Figure S15. Time-dependent decrease of Serum TGF-β Values in RJX-Treated Severe COVID-19 Patients.** [A] Serum **TGF-β** concentration is depicted over 46 days for 4 evaluable responders [B] The first order kinetics of the reductions in serum **TGF-β** values was investigated by fitting a straight line to a semi-log plot of the portion of the **TGF-β** concentration x time curve that displayed maximum reduction in **TGF-β** values over the course of RJX treatment. There were 33 independent data points across 45 days. The slope of the line represents the rate constant for **TGF-β** reduction in $\log_{10}$ scale (viz.: -0.0055). The time to 50% reduction ($T_{50}$ = 55.5 days) of serum **TGF-β** values was calculated using the rate constant and the intercept value of 2.01. Equation of the line: $\text{Log}_{10}$ (% Reduction) = -0.00553 x Time(Days) + 2.01.

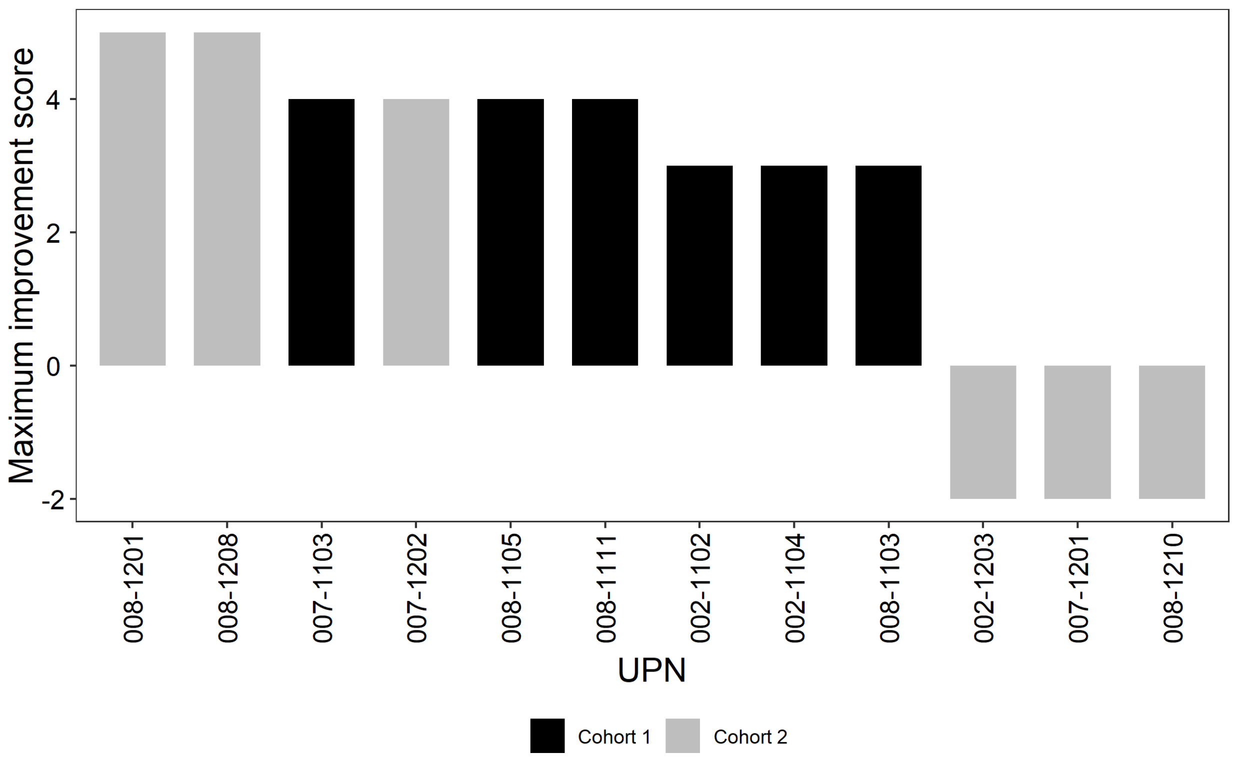


**Figure S16. A Waterfall plot of Maximum Clinical Score Change on 8-Point Ordinal Scale.**

### SUPPLEMENTAL TABLES

#### Table S1: Quantitative Composition of RJX

| Vial | Component | % Content | Content mg/10 mL |
|---|---|---|---|
| Vial A | **API (designated by numbers)** | | |
| | 1) Ascorbic Acid USP | 8.9933 | 899.33 |
| | 2) Thiamine HCl USP | 0.6333 | 63.33 |
| | 3) Magnesium Sulfate Heptahydrate USP | 8.080 | 808.00 |
| | 4) Cyanocobalamin Crystalline USP | 0.0193 | 1.93 |
| | 5) Niacinamide USP | 1.188 | 118.80 |
| | 6) Pyridoxine HCl USP | 1.188 | 118.80 |
| | 7) Riboflavin 5'Phosphate USP | 0.0253 | 2.53 |
| | 8) Calcium D-Pantothenate USP | 0.0293 | 2.93 |
| | Water for Injection USP (diluent) | 79.8425 | 7984.25 |
| | Sodium Chloride USP | 0.001 | 0.1 |
| Vial B | Sodium Bicarbonate USP | 8.40 | 840.00 |
| | Water for Injection USP (diluent) | 91.599 | 9159.90 |
| | Sodium Chloride USP | 0.001 | 0.1 |

Abbreviations: USP = United States Pharmacopeia, API = Active Pharmaceutical Ingredient

This table is included to reader convenience; it was published as Table S1 in Front. Pharmacol. 11, 594321; 10.3389/fphar.2020.594321 (Uckun F. M., Carlson J., Orhan C., Powell J., Pizzimenti N. M., Van Wyk H., et al. Rejuveinix shows a favorable clinical safety profile in human subjects and exhibits potent preclinical protective activity in the Lipopolysaccharide-galactosamine Mouse model of acute respiratory distress syndrome and multi-organ failure)

#### Table S2. Patient Characteristics and Demographic Features for Part 1 of RPI015 Study

| UPN | CH# | Co-morbidities | Height (in cm)/Weight (in kg)/ BMI (in kg/m$^2$) | Hx of Vac./ Covid-19 Dx. | 8-point Ordinal Scale[c] | Pneumonia Findings CxR/CT | CRP mg/L | ALC ($10^9$/L) | Fever/Cough/ SOB/Hypoxia** | Dx-ICF |
|---|---|---|---|---|---|---|---|---|---|---|
| 002-1102 | 1 | Obesity | 167.6/102.8/36.6 | -/+[a] | 4 | + | 64.3 | 0.9 | +/+/+/+ | 8 |
| 002-1203 | 2 | Obesity | 170.2/93.0/32.1 | -/+[a] | 3 | + | 152 | 0.5 | +/+/+/+ | 1 |
| 002-1104 | 1 | Obesity, HTN,DM,HC | 172.7/104.8/35.1 | +/+[a] | 4 | + | 55.3 | 1.4 | -/+/+/+ | 2 |
| 008-1201 | 2 | - | 167.6/66.7/23.8 | -/+[b] | 3 | + | 80.3 | 0.4 | +/+/+/+ | 11 |
| 008-1103 | 1 | Obesity, HTN,Asthma | 157.5/90.7/36.6 | -/+[a] | 4 | + | 94.6 | 0.5 | +/+/+/+ | 10 |
| *008-1204 (withdrew consent on D4)* | 2 | Obesity | 167.7/104.0/37.0 | -/+[a] | 3 | + | 297.6 | 0.5 | +/+/+/+ | 6 |
| 008-1105 | 1 | Obesity, HTN, DM | 177.8/156.0/49.4 | -/+[a] | 4 | + | 106.5 | 0.3 | +/+/+/+ | 7 |
| 008-1208 | 2 | Obesity, CHF, COPD | 162.6/89.4/33.8 | -/+[a] | 3 | + | 42.4 | 1.2 | +/+/+/+ | 4 |
| 008-1210 | 2 | Obesity Asthma | 167.6/115.0/40.9 | +/+[a] | 3 | + | 54.1 | 0.9 | +/+/+/+ | 6 |
| 007-1201 | 2 | Overweight, HTN | 165.1/77.1/28.3 | -/+[a] | 3 | + | 50.3 | 0.5 | -/+/+/+ | 1 |
| 007-1202 | 2 | Obesity, HTN,DM | 152.4/77.1/33.2 | -/+[a] | 3 | + | 54.1 | 0.9 | -/-/+/+ | 0 |
| 008-1111 | 1 | Obesity | 160.0/118/46.1 | -/+[a] | 4 | + | 53.4 | 0.9 | +/+/+/+ | 2 |
| 007-1103 | 1 | Overweight | 149.9/65.8/29.3 | -/+[a] | 4 | + | 87.9 | 0.9 | -/+/+/+ | 1 |

All Cohort 2 patients had HRF requiring high-flow oxygen +/- non-invasive positive pressure ventilation (NIPPV). No patient on invasive mechanical ventilation (IMV) was eligible for this study. All Cohort 1 patients had hypoxia in room air requiring supplemental oxygen (not high-flow, NIPVV, or MV). Covid-19: Corona virus disease 2019; CT: Computed tomography; UPN: Unique patient number; CH: Cohort

[a]FDA-approved RT-PCR test [b] FDA-approved antigen test; [c]8-point ordinal scale: 1. Death; 2. Hospitalized, on invasive mechanical ventilation or ECMO; 3. Hospitalized, on non-invasive ventilation or high flow oxygen devices; 4. Hospitalized, requiring supplemental oxygen; 5. Hospitalized, not requiring supplemental oxygen - requiring ongoing medical care (COVID-19 related or otherwise); 6. Hospitalized, not requiring; supplemental oxygen - no longer requires ongoing medical care; 7. Not hospitalized, limitation on activities and/or requiring home oxygen; 8. Not hospitalized, no limitations on activities. HTN: hypertension; DM: diabetes mellitus; HC: hypercholesterolemia; H/L: Hispanic or Latino

**Patients 007-1201, 008-1102, 008-1103, 008-1210 also had diarrhea. Patient 008-1204 withdrew consent on day 4. Body Mass Index (BMI) was calculated using the formula: Weight(kg) / Height(m)$^2$. In accordance with the federal guidelines on the identification, evaluation, and treatment of overweight and obesity in adults released by the National Heart, Lung, and Blood Institute (NHLBI), "overweight" was defined as a BMI value between 25 and 29.9; and "obesity" as a BMI value greater than or equal to 30.

#### Table S3. Concomitant Standard of Care Therapies

| Patient No. | Antiviral Therapy | Antibiotic Therapy | Anticoagulation | Non-steroidal Anti-Inflammatory Therapy | Corticosteroids |
|---|---|---|---|---|---|
| 002-1102 | Remdesivir, Convalescent plasma | Azithromycin, Ceftriaxone | Enoxaparin | - | Dexamethasone |
| 002-1203 | Remdesivir | Doxycycin, Ceftriaxone, Piperacillin-Tazobactam, | Enoxaparin | - | Dexamethasone, Solu-Medrol |
| 002-1104 | Remdesivir | Azithromycin, Ceftriaxone | Enoxaparin, Rivaroxaban | - | Dexamethasone |
| 008-1201 | Remdesivir | Azithromycin, Ceftriaxone | Enoxaparin | - | Dexamethasone, Budesonide |
| 008-1103 | - | Azithromycin, Ceftriaxone | Enoxaparin | - | Dexamethasone |
| 008-1204 | Remdesivir | Azithromycin, Ceftriaxone | Enoxaparin | - | Dexamethasone |
| 007-1103 | Acyclovir, Gancyclovir | Ertapenem | Enoxaparin | - | Solu-Medrol |
| 008-1105 | - | Piperacillin-Tazobactam, Ceftriaxone | Enoxaparin | - | Dexamethasone |
| 008-1208 | Remdesivir | Azithromycin, Ceftriaxone | Enoxaparin | Ibuprofen | Dexamethasone |
| 008-1210 | Remdesivir | Azithromycin, Ceftriaxone, Linezolid, | Enoxaparin | - | Dexamethasone, Budesonide |
| 007-1201 | - | Ertapenem | Enoxaparin | - | Solu-medrol |
| 007-1202 | - | - | Enoxaparin | - | Solu-medrol, Prednisone |
| 008-1111 | - | Azithromycin, Ceftriaxone | Enoxaparin | - | Dexamethasone |

#### Table S4: Listing of all Grade 3-5 AEs by MedDRA PT: All enrolled patients in Part 1 of the RPI015 study

| UPN | CH# | Preferred term | AE term (CTCAE Grade) | SAE | Date of onset (Number of days from ICF) | Date of resolution (Number of days from ICF) | Duration (days) | Relatedness to RJX | Relatedness to Covid-19 | Action Taken with RJX | Outcome | DLT (Yes/No) | SUSAR (Yes/No) | PDC of RJX |
|---|---|---|---|---|---|---|---|---|---|---|---|---|---|---|
| 002-1203 | Cohort 2 | Acute respiratory failure | Worsening acute hypoxemic respiratory failure (5) | Yes | Day 12 | NA | 15 (CODOD) | No | Yes | NA | NR, Fatal | No | No | No |
| 007-1201 | Cohort 2 | Sepsis | Sepsis (3) | No | Day 10 | NA | 14 (CODOD) | No | Yes | NA | NR | No | No | No |
| | | Fibrin D dimer increased | Elevated D-dimer (3) | No | Day 17 | NA | 7 (CODOD) | No | Yes | NA | NR | No | No | No |
| | | Pulmonary embolism | Acute pulmonary embolism right lower lobe (3) | No | Day 19 | NA | 5 (CODOD) | No | Yes | NA | NR | No | No | No |
| | | Cardiac arrest | Cardiac arrest (5) | Yes | Day 23 | NA | 1 (CODOD) | No | Yes | NA | NR, Fatal | No | No | No |
| | | Intestinal ischemia | Mesenteric ischemia (5) | Yes | Day 23 | NA | 1 (CODOD) | No | Yes | NA | NR, Fatal | No | No | No |
| 008-1210 | Cohort 2 | Acute respiratory failure | Worsening of acute hypoxic respiratory failure (5) | Yes | Day 8 | NA | 6 (CODOD) | No | Yes | NA | NR, Fatal | No | No | No |
| | | Multiple organ dysfunction syndrome | Multi organ failure (5) | Yes | Day 13 | NA | 1 (CODOD) | No | Yes | NA | NR, Fatal | No | No | No |

AE: Adverse event; CH: Cohort; Covid-19: Corona virus disease 2019; CODOD: Censored on date of death; CTCAE: Common terminology criteria for adverse events; DLT: Dose limiting toxicity; ICF: Informed consent form; MedDRA: Medical dictionary for regulatory activities; NA: Not applicable; NR: Not recovered/not resolved; PD: Permanent discontinuation; RJX: Rejuveinix; SAE: Serious adverse event; SUSAR: Suspected unexpected serious adverse reaction; UPN: Unique patient number

#### Table S5: Listing of all grade SAEs by MedDRA PT - All enrolled patients in Part 1 of the RPI015 study

| UPN | CH# | Preferred term | AE term (CTCAE Grade) | Date of onset (Number of days from ICF) | Date of resolution (Number of days from ICF) | Duration (days) | Relatedness to RJX | Relatedness to Covid-19 | Action Taken with RJX | Outcome | DLT | SUSAR | PDC of RJX |
|---|---|---|---|---|---|---|---|---|---|---|---|---|---|
| 002-1203 | Cohort 2 | Acute respiratory failure | Worsening Acute Hypoxemic Respiratory Failure (5) | Day 12 | NA | 15 (CODOD) | No | Yes | NA | NR, Fatal | No | No | No |
| 007-1201 | Cohort 2 | Cardiac arrest | Cardiac Arrest (5) | Day 23 | NA | 1 (CODOD) | No | Yes | NA | NR, Fatal | No | No | No |
| | | Intestinal ischemia | Mesenteric Ischemia (5) | Day 23 | NA | 1 (CODOD) | No | Yes | NA | NR, Fatal | No | No | No |
| 008-1210 | Cohort 2 | Acute respiratory failure | Worsening of acute hypoxic respiratory failure (5) | Day 8 | NA | 6 (CODOD) | No | Yes | NA | NR, Fatal | No | No | No |
| | | Multiple organ dysfunction syndrome | Multi Organ Failure (5) | Day 13 | NA | 1 (CODOD) | No | Yes | NA | NR, Fatal | No | No | No |

AE: Adverse event; CH: Cohort; Covid-19: Corona virus disease 2019; CODOD: Censored on date of death; CTCAE: Common terminology criteria for adverse events; DLT: Dose limiting toxicity; ICF: Informed consent form; MedDRA: Medical dictionary for regulatory activities; NA: Not applicable; NR: not recovered/not resolved; PD: Permanent discontinuation; RJX: Rejuveinix; SAE: Serious adverse event; SUSAR: Suspected unexpected serious adverse reaction; UPN: Unique patient number

#### Table S6: Incidence of Grade 3-5 AEs and SAEs by MedDRA PT and Listing of Deaths: All enrolled patients in Part 1 of the RPI015 study

##### A. Incidence of all Grade 3-5 AE

| MedDRA PT | Cohorts: Cohort 1 (N=6) | Cohort 2 (N=7) | Total: N = 13 n (%) |
|---|---|---|---|
| Acute respiratory failure | 0 | 2 (15.4%) | 2 (15.4%) |
| Cardiac arrest | 0 | 1 (7.7%) | 1 (7.7%) |
| Fibrin D dimer increased | 0 | 1 (7.7%) | 1 (7.7%) |
| Intestinal ischaemia | 0 | 1 (7.7%) | 1 (7.7%) |
| Multiple organ dysfunction syndrome | 0 | 1 (7.7%) | 1 (7.7%) |
| Pulmonary embolism | 0 | 1 (7.7%) | 1 (7.7%) |
| Sepsis | 0 | 1 (7.7%) | 1 (7.7%) |

##### B. Incidence of all SAEs

| MedDRA PT | Cohorts: Cohort 1 (N=6) | Cohort 2 (N=7) | Total: N = 13 n (%) |
|---|---|---|---|
| Acute respiratory failure | 0 | 2 (15.4%) | 2 (15.4%) |
| Cardiac arrest | 0 | 1 (7.7%) | 1 (7.7%) |
| Intestinal ischaemia | 0 | 1 (7.7%) | 1 (7.7%) |
| Multiple organ dysfunction syndrome | 0 | 1 (7.7%) | 1 (7.7%) |

##### C. Listing of Deaths

| D. Patient No. | Cohort# | Timing (Days) from First and Last RJX Infusion | Cause of death | Relatedness to RJX | Relatedness to COVID-19 |
|---|---|---|---|---|---|
| 002-1203 | Cohort 2 | 25 / 19 | SAE: Worsening Acute Hypoxemic Respiratory Failure | No | Yes |
| 007-1201 | Cohort 2 | 21 / 15 | SAE: Cardiac Arrest and Mesenteric Ischemia | No | Yes |
| 008-1210 | Cohort 2 | 14 / 8 | SAE: Worsening Acute Hypoxemic Respiratory Failure ; Multi-Organ Failure | No | Yes |

RJX: Rejuveinix; SAE: Serious adverse event; MedDRA: Medical dictionary for regulatory activities; N: Total number of patients; n: Number of patients with event; PT: Preferred term
